# Leveraging Skin Mutagenesis to Uncover Genetic Cancer Susceptibilities

**DOI:** 10.64898/2026.10.02.26364350

**Authors:** Audris Chiang, Gayathri Swaminathan, Gaurav Luthria, Marlayna Harris, Haniyah Shareef, Shufeng Li, Vivian J. Hua, Warren H. Chan, Hyunje G. Cho, Michelle Jade Lin, Kiana Khalili, Jeanie Ramos, Hanh Do, Mahrukh Abdullah, Irene Bailey, 23andMe Research Team, Kerri E. Rieger, Jiali Han, Paul J. Utz, Christina Curtis, Anthony E. Oro, Kavita Y. Sarin

**Affiliations:** Department of Dermatology, Stanford University School of Medicine; Stanford, CA, USA; Department of Dermatology, Icahn School of Medicine at Mount Sinai; New York, NY, USA; Department of Pediatrics, University of California Davis School of Medicine; Sacramento, CA, USA; 23andMe, Inc.; South San Francisco, CA, USA; Department of Pathology, Stanford University School of Medicine; Stanford, CA, USA; Department of Epidemiology, Indiana University Richard M. Fairbanks School of Public Health; Indianapolis, IN, USA; Indiana University Melvin and Bren Simon Comprehensive Cancer Center; Indianapolis, IN, USA; Department of Medicine, Immunology and Rheumatology, Stanford University School of Medicine; Stanford, CA, USA; Department of Medicine, Genetics and Biomedical Data Science, Stanford University School of Medicine; Stanford, CA, USA

## Abstract

Identifying inherited cancer predisposition genes remains challenging. We leveraged the skin’s high UV-mutagenic burden to discover susceptibility genes by studying patients with high frequency of basal cell carcinomas (hfBCC). We found that hfBCC patients had three-fold higher internal malignancy prevalence and identified a novel candidate *EPHB2* cancer susceptibility mutation associated with BCC and prostate cancer. These findings demonstrate the skin’s potential as a powerful platform for discovering cancer predisposition variants.

---

5-10% of cancers arise in individuals with inherited cancer predisposition.^1^ Identifying these genetically-predisposed individuals can inform risk assessment, surveillance, and early cancer detection.^2^ While some cancer susceptibility genes are known, many more remain undiscovered, highlighting the need for novel approaches to identify cancer susceptibility genes.^3^

Ultraviolet (UV) exposed skin experiences sustained mutagenic stress and depends on intact DNA-repair, immune surveillance, and checkpoint pathways to prevent malignant transformation.^4,5^ Basal cell carcinoma (BCC), the most common skin cancer caused by UV exposure, provides a visible readout of how inherited cancer-protective mechanisms perform under repeated environmental injury. We hypothesized that patients who develop unusually high numbers of BCCs represent an extreme phenotype enriched for inherited defects in cancer-protective pathways. Supporting this hypothesis, we previously identified an increased prevalence of germline DNA-repair mutations among patients with high-frequency BCC (hfBCC).^6^ These observations suggest that hfBCC may serve as a marker of inherited cancer susceptibility, highlighting the potential of our skin to serve as a sentinel organ in identifying new genes that predispose to cancer.

We evaluated this extreme phenotype strategy by integrating whole-genome sequencing (WGS), internal cancer epidemiology, population-scale association, and functional studies. We sought to determine if hfBCC identified a recognizable population with a higher prevalence of internal malignancy and whether this phenotype could facilitate discovery of novel germline cancer susceptibility variants.

We enrolled 94 individuals with hfBCC, defined as ≥6 biopsy-proven BCCs within a 10-year time period (63% male; mean age at first diagnosis 41 years; mean BCC count 17 [range 6-98]), and 136 participants with low numbers of BCC in an age-matched control cohort (43% male; mean age 63 years; mean lifetime BCC count 0.21) (**Table S1**). Twenty-four participants with hfBCC (26%) had a history of internal malignancy, corresponding to a 3.3-fold higher prevalence than age-adjusted Surveillance, Epidemiology, and End Results (SEER) population estimates (Relative Risk 3.3, 95% confidence interval (CI) 2.5-4.3; p<0.0001, χ^2^ test) (**Table 1**). This association was independently corroborated in the Merative™ MarketScan^®^ Commercial Database (DOI: <u>10.57761/p0ta-q619</u>), a US insurance claims database representing over 250 million individuals (Odds Ratio 1.9, 95% CI 1.8-2.1; p<0.0001, two-sided multivariable logistic regression adjusting for age and sex) (**Table S2**). These results confirm that hfBCC patients have increased risk for internal cancers, and the development of hfBCC can be a marker to identify patients harboring cancer susceptibility.

**Table 1.** Internal Cancer Prevalence in hfBCC Cohort (N = 94). Table of cancer prevalence of hfBCC cohort compared to expected prevalence derived from age-adjusted prevalence proportions in the Surveillance Research Program, National Cancer Institute SEER database. SEER reference population ages 50 to 84 years old were included (N=45,256,676). Statistical analysis was conducted using Fisher’s exact test for analyses with number hfBCC cancer cases ≤5 and Chi-square test otherwise to compare observed cancer counts in the hfBCC cohort and expected counts from SEER prevalence. RR = relative risk. CI = confidence interval.

| Cancer | Number cases in |  |  |  |
| --- | --- | --- | --- | --- |
|  | hfBCC Cohort<br>(%) | Number SEER cases<br>(%)<sup>a</sup> | RR in hfBCC vs SEER (95%<br>CI) | P value |
| Leukemia / Lymphoma | 8 (8.5) | 403,759 (0.9) | 9.54 (4.90 – 17.83) | <0.0001 |
| Colon | 2 (2.1) | 256,470 (0.6) | 3.75 (1.03 – 13.11) | 0.0999 |
| Breast<sup>b,c</sup> | 4 (11.4) | 1,057,308 (4.4) | 2.55 (1.01 – 5.79) | 0.0702 |
| Ovarian<sup>b,c</sup> | 1 (2.9) | 52,292 (0.2) | 12.88 (2.28 – 65.54) | 0.0748 |
| Parotid | 2 (2.1) | 13,991 (0.03) | 68.82 (18.93 – 240.30) | 0.0004 |
| Prostate<sup>b,c</sup> | 11 (18.6) | 1,054,131 (5.2) | 3.83 (2.21 – 6.25) | <0.0001 |
| Renal | 2 (2.1) | 205,528 (0.5) | 4.69 (1.29 – 16.36) | 0.0685 |
| Any Solid Organ Malignancy<sup>d</sup> | 27 (28.7) | 4,285,890 (9.5) | 3.03 (2.17 – 4.07) | <0.0001 |
| Any Internal Malignancy<sup>d</sup> | 32 (34.0) | 4,627,559 (10.3) | 3.33 (2.47 – 4.31) | <0.0001 |

<sup>a</sup>SEER counts are based on age-adjusted prevalence proportions derived using SEER\*Stat software (v9.0.42.0).
<sup>b</sup>Ovarian and breast cancer are calculated among females. Prostate cancer is calculated among males.
<sup>c</sup>The SEER population used for sex-specific cancers are female (N = 23,581,900) or male (N = 21,674,776).
<sup>d</sup>Other history of internal cancers include pancreatic, lung, throat, and bladder cancers.

Germline WGS identified seven recurrent enriched germline variants (**Figure 1a**; **Table 1; Table S3**). The leading candidate was recurrent hotspot variant *EPHB2* p.D679N (G2035A, dbSNP rs28936395), detected in five participants with hfBCC and enriched five-fold relative to the gnomAD Non-Finnish European (NFE) reference population (p<0.001, χ^2^ test), and seven-fold relative to our age-matched control cohort (p=0.044, two-sided Fisher’s exact test). Four of five carriers had a history of internal malignancy, and all 3 male carriers had prostate cancer (p=0.024, two-sided Fisher’s exact test, relative to SEER estimates) (**Table 3**). Single-cell RNA sequencing (scRNA-seq) demonstrated preferential *EPHB2* expression in BCC keratinocytes (**Figure 1b; Figure S1**), with immunohistochemistry supporting EphB2 expression in BCC keratinocytes (**Figure S2**). The p.D679N variant occurs within the EphB2 kinase domain (**Figure 1c; Figure S3**).

**Figure 1.**
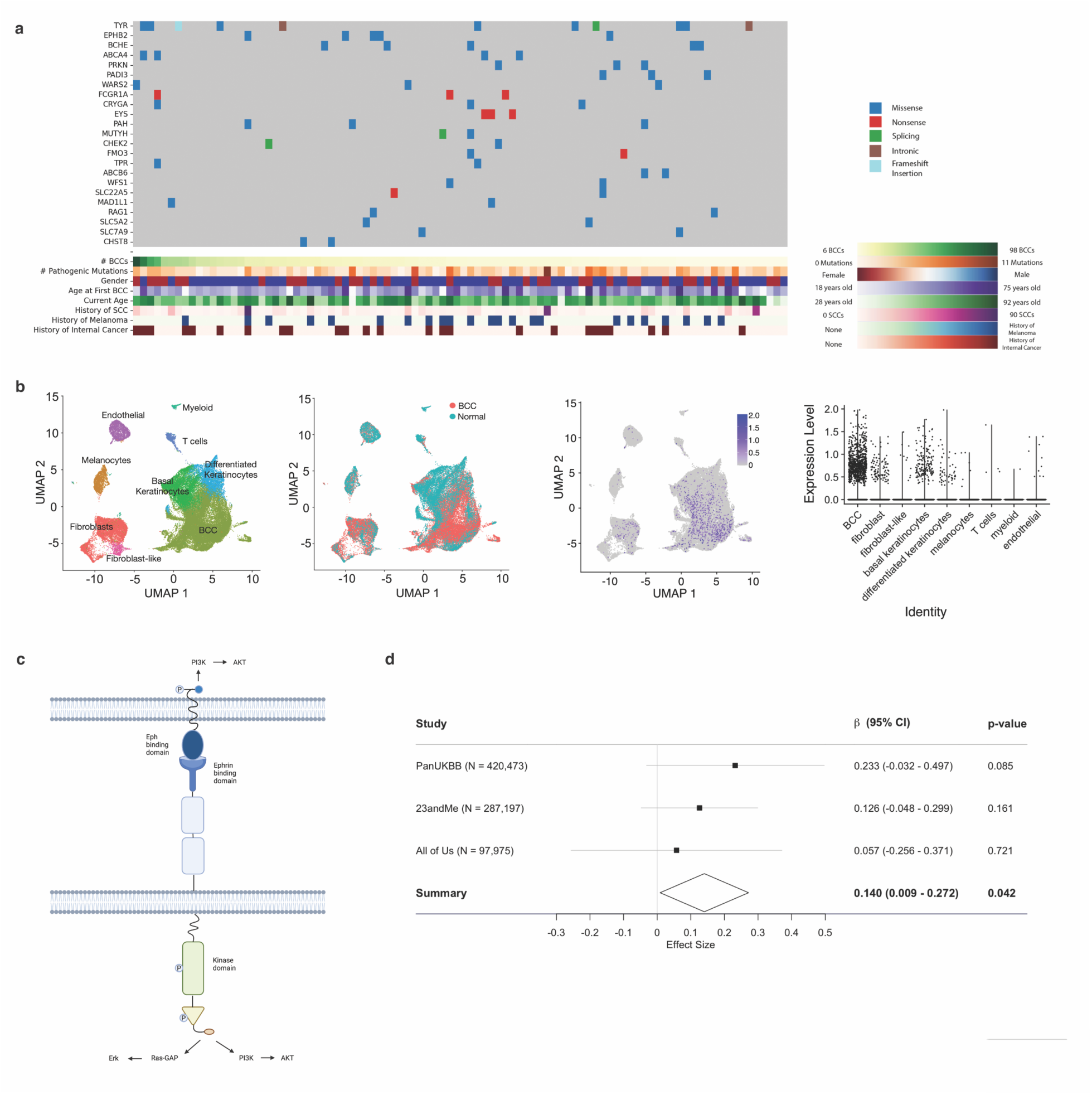
High-frequency BCC is associated with higher internal malignancy prevalence and identifies the recurrent germline hotspot variant *EPHB2* p.D679N. (**a**) Heatmap of recurrent pathogenic germline variants identified in hfBCC cohort. (**b**) 2D UMAP visualization of scRNA-seq data with single cells clustered by cell-types, color-coded by sample type. *EPHB2* expression clustered primarily in BCC cells demonstrated in UMAP and violin plots. (**c**) *EPHB2* encodes receptor tyrosine kinase EphB2, which interfaces with various pathways including PI3K/Akt and MAPK/ERK, and leads to context-dependent upregulation or downregulation of cell proliferation and growth. (**d**) Forest plot of GWAS meta-analysis of *EPHB2* p.D679N provides evidence of enrichment in BCC patients (≥ 1 BCCs) within Pan-UK Biobank, 23andMe, and All of Us cohorts (p = 0.042). Error bars = 95% confidence intervals. Diamond = pooled effect.

To assess *EPHB2* p.D679N in the broader BCC phenotype, we performed meta-analysis across 20,219 individuals with ≥1 BCC and 785,426 controls without BCCs from Pan-UK Biobank, 23andMe, Inc., and All of Us genome-wide association study (GWAS) cohorts. Although no individual cohort reached statistical significance, pooled meta-analysis provided support for association of *EPHB2* p.D679N with BCC (β=0.140, SE=0.067; p=0.042, PLINK inverse-variance weighted fixed effects model) (**Figure 1d**). Meta-analyses of the other six enriched variants were not significant (**Figure S4**).

In post hoc comparison, p.D679N demonstrated 4.67-fold enrichment in extreme phenotype hfBCC versus 1.15-fold enrichment in conventional BCC, representing approximately 4-fold greater relative enrichment with extreme-phenotype ascertainment. Under a standardized model assuming a dominant carrier effect, 1:1 case-control allocation, 80% power, and two-sided α = 0.05, this difference corresponded to an estimated 230-fold improvement in case-sample efficiency.

To evaluate the signaling effects of *EPHB2* p.D679N, mutant EPHB2 was expressed in human BCC cell lines and compared with wild-type EPHB2 and vector control. The variant did not alter receptor autophosphorylation (**Figure 2a; Figure S5a**), but reduced EphB2 protein abundance by 57% (p=0.003, two-tailed Student’s *t*-test) and increased ERK1/2 phosphorylation 1.5-fold (p=0.023, two-tailed Student’s *t*-test), without significantly increasing Akt phosphorylation (p=0.154, two-tailed Student’s *t*-test) (**Figure 2b; Figures S5b and S6**). Consistent with the role of EphB2 in modulating MAPK signaling, these findings suggest that the D679N variant alters downstream pathway activity rather than acting as a classical activating kinase mutation.^7,8^

**Figure 2.**
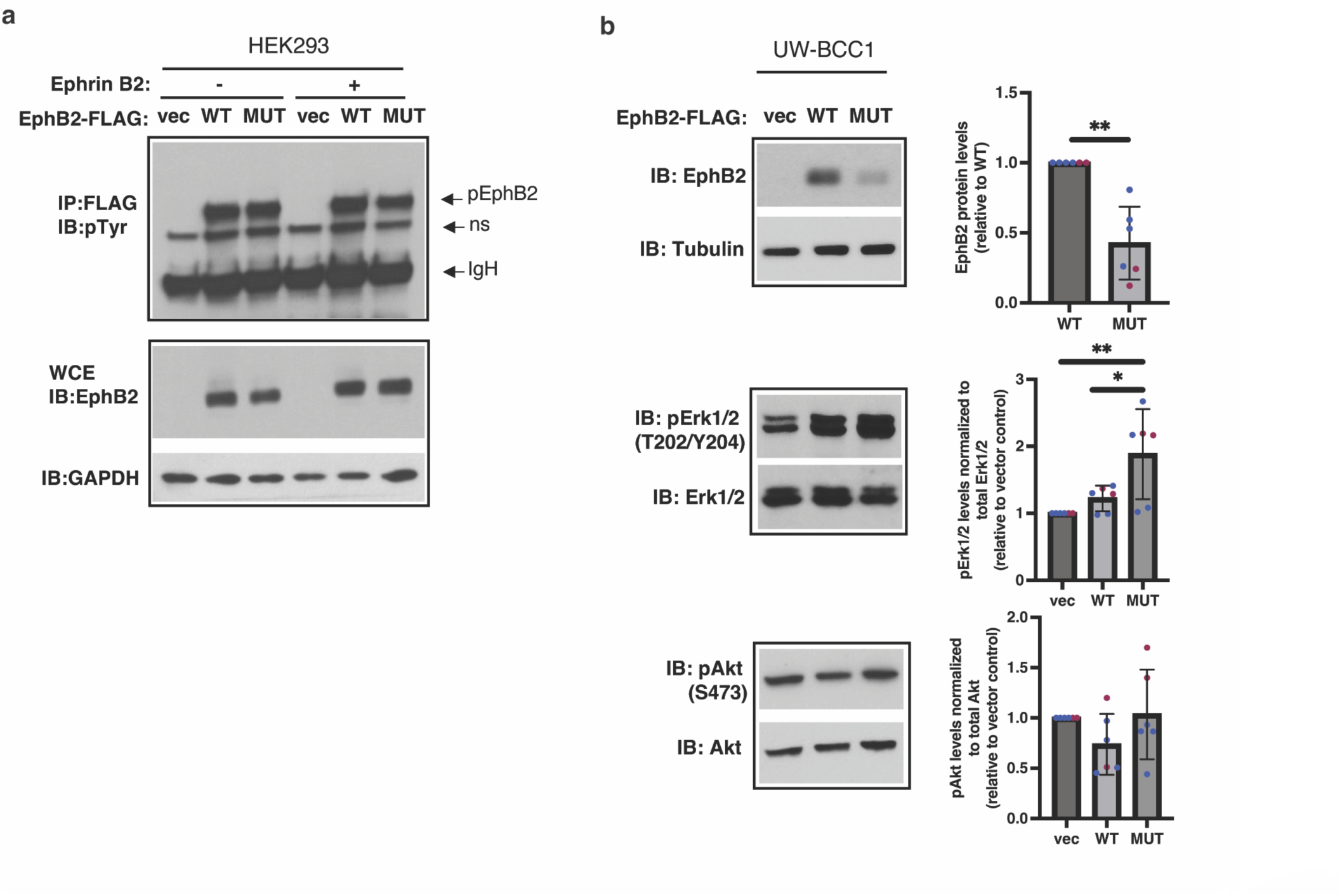
Signaling effects of the *EPHB2* p.D679N variant. (**a**) 48h post-transfection of HEK293 cells with FLAG-tagged wild-type (WT), D679N mutant (MUT), or vector (vec) control, with (+) and without (-) Ephrin B2-Fc (1.5 mg/ml) 15 min stimulation. Tyrosine phosphorylation of WT versus MUT EphB2, assessed by immunoblotting of FLAG immunoprecipitates with a phospho-tyrosine antibody (4G10), were not significantly different. ns = non-specific, IgH = Ig heavy chain. (**b**) Levels of WT and MUT EphB2, phosphorylated and total Erk, and phosphorylated and total Akt, were evaluated in UW-BCC1 (blue) and UW-BCC1-T2 (violet) cell lines transfected with WT, MUT, or vec. Representative blots and quantitation of EphB2, pAkt, and pErk normalized to Tubulin and GAPDH, Akt, and Erk, respectively, are shown for UW-BCC1 cell line. MUT EphB2 expression was significantly lower than WT (p = 0.003), and MUT pErk upregulation significantly higher than WT (p=0.023). MUT pAkt upregulation did not reach significance. *p < 0.05, **p < 0.01.

Overall, our findings demonstrate that individuals with hfBCC have a higher prevalence of internal malignancy, supporting the hypothesis that extreme BCC burden identifies a recognizable subgroup enriched for inherited cancer susceptibility. Because the skin experiences lifelong UV-induced mutagenic stress and depends on intact cancer-protective pathways, it may function as a sentinel organ for underlying deficiencies in cancer protection. A history of numerous BCCs may therefore represent a useful clinical feature that can inform referral for hereditary cancer risk assessment, although prospective validation is needed before hfBCC is incorporated into clinical decision-making.

*EPHB2* p.D679N emerged as a leading candidate cancer susceptibility variant, supported by enrichment in the hfBCC cohort, population-level evidence in BCC meta-analysis, and altered EphB2 abundance and ERK signaling in human BCC models. *EPHB2* encodes EphB2, a receptor tyrosine kinase involved in regulating cell proliferation, migration, and metabolism with context-dependent tumor-promoting and tumor-suppressive functions.^9–11^ The observation of prostate cancer in all three male carriers was notable, suggesting potential relevance of the germline variant to prostate cancer. Although *EPHB2* p.D679N was previously reported as a somatic variant in prostate and colorectal cancers,^12,13^ its germline cancer associations had not been established. Consistent with known EphB2 signaling biology,^7,8^ the variant reduced EphB2 protein abundance and increased ERK activation, supporting altered MAPK signaling as a potential mechanism contributing to cancer susceptibility, which requires validation in endogenous models.

Our study provides proof-of-principle that studying individuals with extreme cancer phenotypes, such as those with high numbers of BCCs, can enrich germline cancer susceptibility discovery. Selecting 94 individuals by an extreme cancer phenotype, hfBCC, concentrated a rare candidate variant signal that received population-level support only after pooling 20,219 BCC cases and 785,426 controls. Post hoc comparison estimates that this extreme phenotype-informed recruitment reduced case numbers needed by approximately 230-fold in comparison to conventional phenotype ascertainment. The marked difference in discovery cohort size highlights the potential of clinically recognizable extreme phenotypes to accelerate the discovery of rare candidate cancer susceptibility variants.

Several limitations warrant consideration. The rare nature of hfBCC constrained participant recruitment and resulted in a smaller cohort size. The rarity of hfBCC has similarly contributed to the absence of an independently assembled hfBCC cohort available for validation in a comparable population. In addition, genetic analyses were largely restricted to individuals of European ancestry, reflecting the higher prevalence of UV-associated keratinocyte cancers in this population, but potentially limiting the generalizability of our findings. Accordingly, further research is needed to translate these findings into clinical practice.

In conclusion, high-frequency BCC was associated with a higher prevalence of internal malignancy in two retrospective datasets and identified an enriched *EPHB2* variant with population-level support and altered signaling in BCC models. These findings nominate *EPHB2* p.D679N as a candidate cancer susceptibility variant and illustrate the potential of clinically defined extreme phenotypes for germline susceptibility discovery.

## MATERIALS AND METHODS

### Experimental Design

The objective of this study was to identify rare germline variants that confer susceptibility to cancer by leveraging the skin’s unique mutagenic environment. We employed a novel observational mutagenesis screen using an extreme phenotype cohort, individuals with six or more basal cell carcinomas (BCCs) over a 10-year period (high-frequency BCC, hfBCC), as a discovery strategy for inherited defects in cancer protection pathways.

Individuals with hfBCC but without known PTCH1 or SUFU mutations (excluding basal cell nevus syndrome), were enrolled in this study and compared them to a control cohort with minimal BCC burden (0–1 BCCs in a lifetime). Germline whole genome sequencing (WGS) was performed on saliva samples from both cohorts to identify rare variants enriched in the hfBCC group. Variant frequencies were further compared to the gnomAD Non-Finnish European reference population.

To assess the biological relevance of candidate genes, we integrated single-cell RNA sequencing (scRNA-seq) data from BCC tumors and matched peri-tumoral skin, focusing on gene expression within BCC and basal keratinocyte populations. Protein expression of the gene with highest expression in BCCs from our scRNA-seq analysis, *EPHB2*, was examined by immunohistochemistry to identify increased EphB2 protein expression within BCC and basal keratinocytes.

We then evaluated broader associations through candidate gene meta-analysis across three large GWAS datasets (UK Biobank, 23andMe, and All of Us), assessing the relationship between identified variants and BCC risk in the general population. Functional validation was performed in vitro using BCC and other relevant cell lines to assess protein expression and downstream signaling.

This multi-tiered approach, combining extreme phenotype-based germline WGS with transcriptomic profiling, population-level genetic validation, and functional characterization, provides a strategy to uncover inherited cancer susceptibility variants.

### Ethics Declaration

This study was reviewed and approved by the Stanford University Institutional Review Board (IRB protocol 36263). All participants enrolled in the hfBCC cohort and control cohort provided written informed consent after the nature and possible consequences of the study were explained prior to study participation.

Analyses of deidentified data from previously published or publicly available cohorts were conducted in accordance with the ethical approvals and informed consent procedures of the original studies.

### Sample Collection

94 patients without Basal Cell Nevus Syndrome who developed a high frequency of BCCs were enrolled in our study. hfBCC patients were defined as those with six or more BCCs in a ten-year time frame. The majority (60 patients) were seen at Stanford Hospital and Clinics and identified to meet enrollment criteria via chart review and pathology records, out of 2,407 patients seen for a new biopsy-confirmed BCC at Stanford Hospital and Clinics. The remaining 34 patients were enrolled in our study following referral from outside dermatologists, with enrollment contingent upon written and/or pathological proof of a minimum of six or more BCCs in a ten-year time frame. Saliva samples were collected from the 94 patients for whole genome sequencing and analysis. Clinical surveys were completed by 90 of the participants. Basal cell nevus syndrome patients were excluded based on chart review, and identification of germline pathogenic *PTCH1* or *SUFU* mutations with germline whole genome sequencing.

136 patients with criteria of having 0 or 1 BCC in their lifetime based on their self-reported personal history and chart review were enrolled as a comparison control cohort of patients with low number of BCCs (average 0.21 BCCs). Saliva samples were collected from the 136 patients for whole genome sequencing and analysis similar to the hfBCC cohort. The control cohort average age was 63 years-old, with 43% male and 57% female.

Analysis of cell-type specific expression for selected genes was conducted based on single-cell RNA sequencing (scRNA-seq) data obtained from 6 surgically discarded tissues (basal cell carcinoma (BCC), n = 4; peri-tumor skin (PTS), n = 2) obtained from excisional human biopsy specimens at Stanford University Hospital (Palo Alto, California, USA). BCCs were classified into superficial, nodular and infiltrative BCC (ID: BCC-I); superficial and nodular BCC (ID: BCC-II); unknown/“hybrid” (ID: BCC-III); and infiltrative with perineural invasion BCC (ID: BCC-IV) subtypes by board-certified dermatopathologist. Consent was waived given Stanford University institutional approval for use of previously collected de-identified tissue (IRB 29381).

### Surveillance, Epidemiology, and End Results-estimated (SEER-estimated) Analysis

Cancer prevalence was based on age-adjusted prevalence proportions derived using the Surveillance Research Program, National Cancer Institute SEER*Stat software (version 9.0.42.0) from the SEERStat Database: Incidence – SEER Research Limited-Field Data, 21 Registries (excluding Illinois).^14^ The reference population used was of European descent, both sexes, and 50 to 84 years old (N=45,256,676), except for sex-specific cancers (breast, ovarian, prostate cancers) for which reference population were for females (N=23,581,900) or males (N=21,674,776). Comparisons between observed cancer counts in the hfBCC cohort and expected counts derived from SEER prevalence estimates were performed using Fisher’s exact test for analyses with number hfBCC cancer cases ≤5 and Chi-square test otherwise using Prism 10 software.

### Merative™ MarketScan^®^ Commercial Database Dataset Selection and Analysis

The Merative™ MarketScan^®^ Commercial Database represents claims data from approximately 100 commercial plans, representing over 250 million individuals in all US census regions from 2007 onward. No identifiable protected health information is contained in this data set. Individuals age 60 years or older during period 2007-2022 with minimum 10 years enrollment time were identified. A diagnosis of each BCC event was based on a combination of ICD-9 or ICD-10 CM codes and CPT codes associated with destruction, excision, or Mohs procedure (Tables S4 and S5). We selected age- and sex-matched controls for each BCC case. Internal malignancies were also determined from ICD-9 or ICD-10 CM codes (Table S4). The BCC cohort identified in the MarketScan^®^ database consisted of 212,475 patients with 1 BCC, 6,101 with 6 or more BCCs, and 669 with 12 or more BCCs, each within a 10-year time period. The control cohort consisted of 3,017,725 patients with no BCCs. The BCC cohort had an average age of 59.2, 63.4, and 63.2 at first enrollment, respectively, and control cohort with average age 55.7 at first enrollment. Logistic regression, adjusting for age and sex, was applied to estimate the associations between BCC groups (BCC 1, 6 or more, and 12 or more, each within 10 years) and control (no BCCs during enrollment) for each cancer.

### Whole Genome Sequencing and Analysis

DNA isolated from saliva was subjected to whole genome sequencing for a minimum 30X read coverage on the DNBSEQ platform (average 40-fold coverage). Sequencing data were mapped to the human reference genome sequence hg19 using the Burrows-Wheeler Aligner (BWA) (version 0.7.17) (RRID:SCR_010910).^15^ Picard tools (version 2.18.11) (RRID:SCR_006525) was used for SAM to BAM conversion and marking of PCR duplicates, and Genome Analysis Toolkit (GATK) 4 (version 4.4.0.0) (RRID:SCR_001876) was used for base quality score recalibration, indel realignment, and duplicate removal prior to variant calling using HaplotypeCaller.^16,17^ Variants were filtered based on variant quality score recalibration, and then annotated with ANNOVAR (version 2020-06-07) (RRID:SCR_012821).^18^

### Mutational Analysis

Variants were prioritized using the following criteria. Synonymous variants, and variants with allele frequency more than 0.01 in ESP6500SI-V2 (RRID:SCR_012761), 1000 Genomes 1000g2015aug_eur (RRID:SCR_008801), Exome Aggregation Consortium (ExAC) Non-Finnish European (NFE) (release 0.3.1) (RRID:SCR_004068), or Genome Aggregation Database (gnomAD) NFE (v2.1) (RRID:SCR_014964) databases were excluded.^19–21^ Inclusion criteria required that variants were classified as “pathogenic” or “likely pathogenic” by either ClinVar (release 20220320) (RRID:SCR_006169) or InterVar (release 20180118), as determined according to the American College of Medical Genetics and Genomics and Association for Molecular Pathology consensus criteria guidelines.^22,23^ Candidate variants were ordered by hfBCC cohort frequency. Variant fold enrichment was calculated by comparison of variant cohort frequency compared to that of reference population variant allele frequencies obtained from gnomAD NFE.^24^ The hfBCC cohort had on average 2.60 germline pathogenic variants per patient, and 95% of the cohort had at least 1 pathogenic variant. Heatmap of variants was created using Python library seaborn v0.11.2 (RRID:SCR_008394).^25^

### Single-cell RNA-sequencing (scRNA-seq) Data Processing and Analysis

Four basal cell carcinoma tumor and two adjacent peri-tumor skin specimens were surgically excised from human donors at Stanford University Hospital (Palo Alto, California, USA) and immediately shipped to University of California, Irvine (Irvine, California, USA) for processing. The details of processing including cell isolation, 3’-droplet-enabled scRNA-seq raw data processing, and doublet/multiplet simulation and low-quality cell pruning methods are fully described in the methods section of our prior publication on scRNA-seq analysis of human BCCs.^26^

Analysis of cell-type specific expression for selected genes was conducted based on the scRNA-seq data with 54,021 single cells. The resulting gene x cell matrix was stored as a Seurat object (Seurat Version 4.1.3) (RRID:SCR_016341),^27–30^ and log-normalized with a scale factor of 10,000. Cell-type identities (basal keratinocytes, differentiated keratinocytes, melanocytes, fibroblasts, fibroblast-like, etc.) were provided from data annotations ^26^. scRNA-seq visualizations were created by projecting the Seurat gene x cell matrix into a 2D UMAP plot using the Seurat FeaturePlot function. For a specific gene of interest, cells with a minimum gene count > 0 were highlighted. Corresponding violin plots for each gene of interest were constructed using the Seurat VlnPlot function with an overlaying dot plot representing single cells. Significance values for each gene of interest were determined using the Seurat FindAllMarkers function utilizing a Wilcoxon Ranksum Test. The minimum fraction of cells that must encode the gene (min.pct) and the log fold change threshold (logfc.threshold) were set to 0 to include all genes of interest. All analyses were performed using R version 4.2.1 (RRID:SCR_001905).

### Immunohistochemistry (IHC)

IHC was performed on a BCC tumor sample to identify wildtype EphB2 protein level expression. Slides were deparaffinized, then blocked using endogenous hydrogen peroxide 0.3% in phosphate buffered saline (PBS) for 15 minutes at room temperature. Antigen retrieval was done using a pressure cooker. Primary antibody human/mouse EphB2 antibody (R&D Systems, Catalog # AF467) (RRID:AB_355375) in diluents (1xPBS, 0.1% bovine serum albumin (BSA), and 0.1% sodium azide) were applied at working concentration 1/50. Secondary antibody horse anti-goat IgG (Vector Laboratories, Catalog # MP-7405) (RRID:AB_2336526) was then applied. Matching no primary antibody control was conducted on the same BCC tumor sample. Slides were reviewed and images extracted using Leica Biosystems Imaging, Inc. Aperio ImageScope v.12.3.3 (RRID:SCR_020993).

### Meta-analysis

Meta-analysis of the *EPHB2* p.D679N variant was conducted across 3 GWAS studies. In Pan-UKBB (GWAS of UK Biobank’s dataset, using GRCh37 reference genome coordinates for variant position, Chr1: Position 23233346), we evaluated 3,947 patients with BCC and 416,526 controls, identifying the variant with beta effect size (β) 0.2327, standard error (SE) 0.1349 (p=0.0849).^31^ In 23andMe, Inc. GWAS dataset (using GRCh37 reference genome coordinates) with 12,945 BCC patients and 274,252 controls, the variant was identified with β 0.1259, SE 0.0885 (p=0.1614).^32^ Using phecodeX CA103.21 basal cell carcinoma in the *All of Us* (AOU) cohort (using GRCh38 reference genome coordinates for variant position, Chr1: Position 22906853), there were 3327 BCC patients and 94,648 control patients without BCC, and the variant was identified with β 0.0573, SE 0.1600 (p=0.7210).^33^ Meta-analysis was significant with β 0.1403, SE=0.0672 (p=0.0419).

In contrast, meta-analyses of additional enriched variants were not significant (Supplemental Figure S4):

BCHE p.A212V rs114706984, GRCh37 Chr3:165548187, GRCh38 Chr3:165830399: Pan-UKBB β 0.0927, SE 0.1555 (p=0.5508); 23andMe β −0.1490, SE 0.1136 (p=0.1964); AOU β −0.3920, SE 0.2010 (p=0.0514). Meta-analysis β −0.1284, SE 0.0835 (p=0.1204).

SPG7 p.A510V rs61755320, GRCh37 Chr16:89613145, GRCh38 Chr16:89546737: Pan-UKBB β 0.0351, SE 0.1259 (p=0.7816); 23andMe β 0.0026, SE 0.0917 (p=0.9773); AOU β 0.4240, SE 0.1570 (p=0.0070). Meta-analysis β 0.1284, SE 0.0670 (p=0.2141).

PRKN p.R275W rs34424986, GRCh37 Chr6:162206852, GRCh38 Chr6:161785820: Pan-UKBB β −0.1171, SE 0.1782 (p=0.5101); 23andMe β 0.0147, SE 0.1492 (p=0.9207); AOU β 0.0234, SE 0.2200 (p=0.9150). Meta-analysis β −0.0262, SE 0.1015 (p=0.8631).

MPEG1 p.T73A rs143527301, GRCh37 Chr11:58980122, GRCh38 Chr11:59212649: Pan-UKBB β −0.2712, SE 0.1871 (p=0.1468); 23andMe β −0.0764, SE 0.1444 (p=0.6002); AOU β −0.1800, SE 0.2060 (p=0.3820). Meta-analysis β −0.1564, SE 0.1000 (p=0.1549).

WARS2 p.W13G rs139548132, GRCh37 Chr1:119683231, GRCh38 Chr1:119140608: Pan-UKBB β 0.1613, SE 0.1937 (p=0.4038); not present in 23andMe GWAS; AOU β 0.0573, SE 0.1890 (p=0.7620). Meta-analysis β 0.1080, SE 0.1353 (p=0.4113).

CRYGA p.R80H rs139353014, GRCh37 Chr2:209027941, GRCh38 Chr2:208163217: Pan-UKBB β −0.0475, SE 0.2032 (p=0.8164); 23andMe β 0.2642, SE 0.1398 (p=0.0669); AOU β −0.1380, SE 0.2180 (p=0.5250). Meta-analysis β 0.0696, SE 0.1018 (p=0.2726).

Meta-analyses were conducted using PLINK v1.90b7 (http://pngu.mgh.harvard.edu/purcell/plink/) (RRID:SCR_001757) and standard errors were calculated using inverse-variance weighting of individual study standard errors.^34^ Forest plots were created using R package forestplot v3.1.3 (https://CRAN.R-project.org/package=forestplot).

### Post hoc comparison of enrichment and estimated case-sample efficiency

We performed a post hoc, exploratory analysis comparing the enrichment and estimated case-sample efficiency of *EPHB2* p.D679N under extreme-phenotype hfBCC ascertainment, defined as selecting cases with ≥6 BCCs within 10 years, versus conventional BCC ascertainment, defined as selecting cases with ≥1 BCC. Among 94 individuals with hfBCC, five heterozygous carriers were identified, corresponding to a carrier frequency of 0.0532 and an allele frequency of 0.0266 [5 / (2*94)]. Relative to gnomAD Non-Finnish European reference allele frequency of 0.0057, this represented 4.67-fold allele frequency enrichment (Table 2).

**Table 2.**
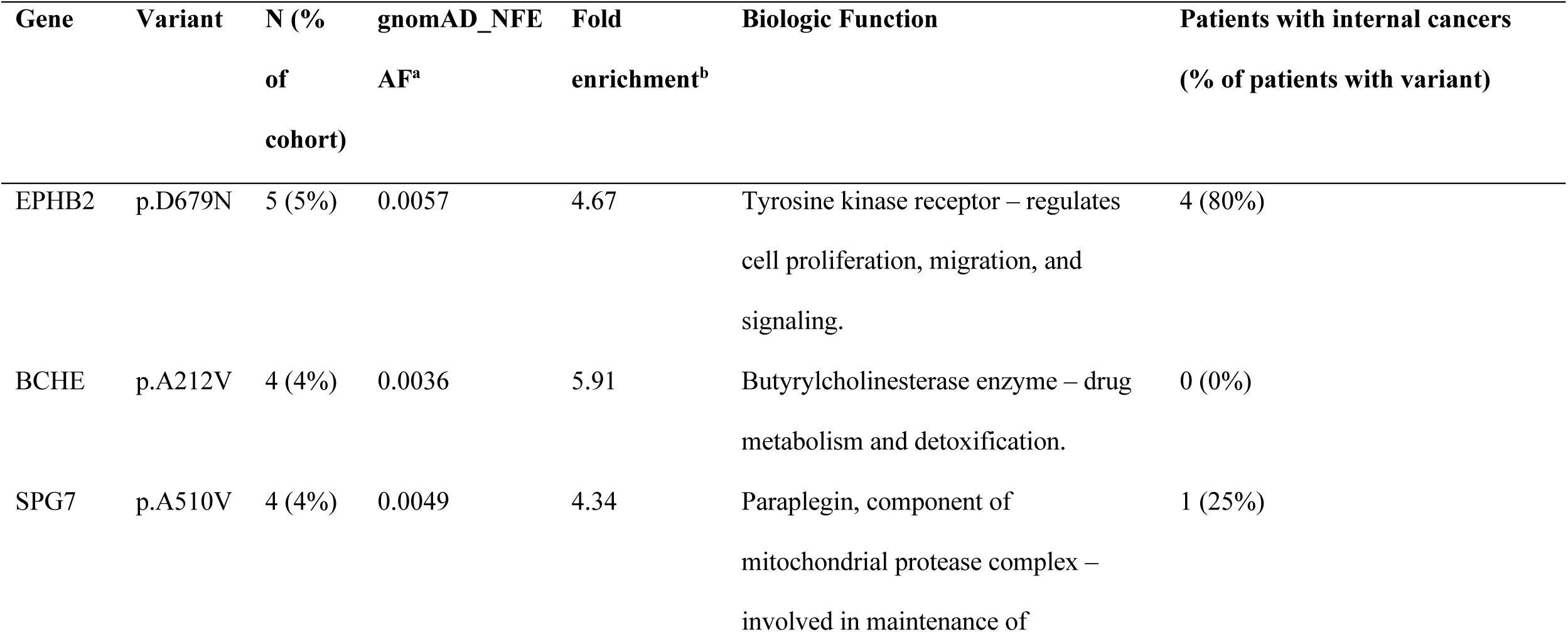
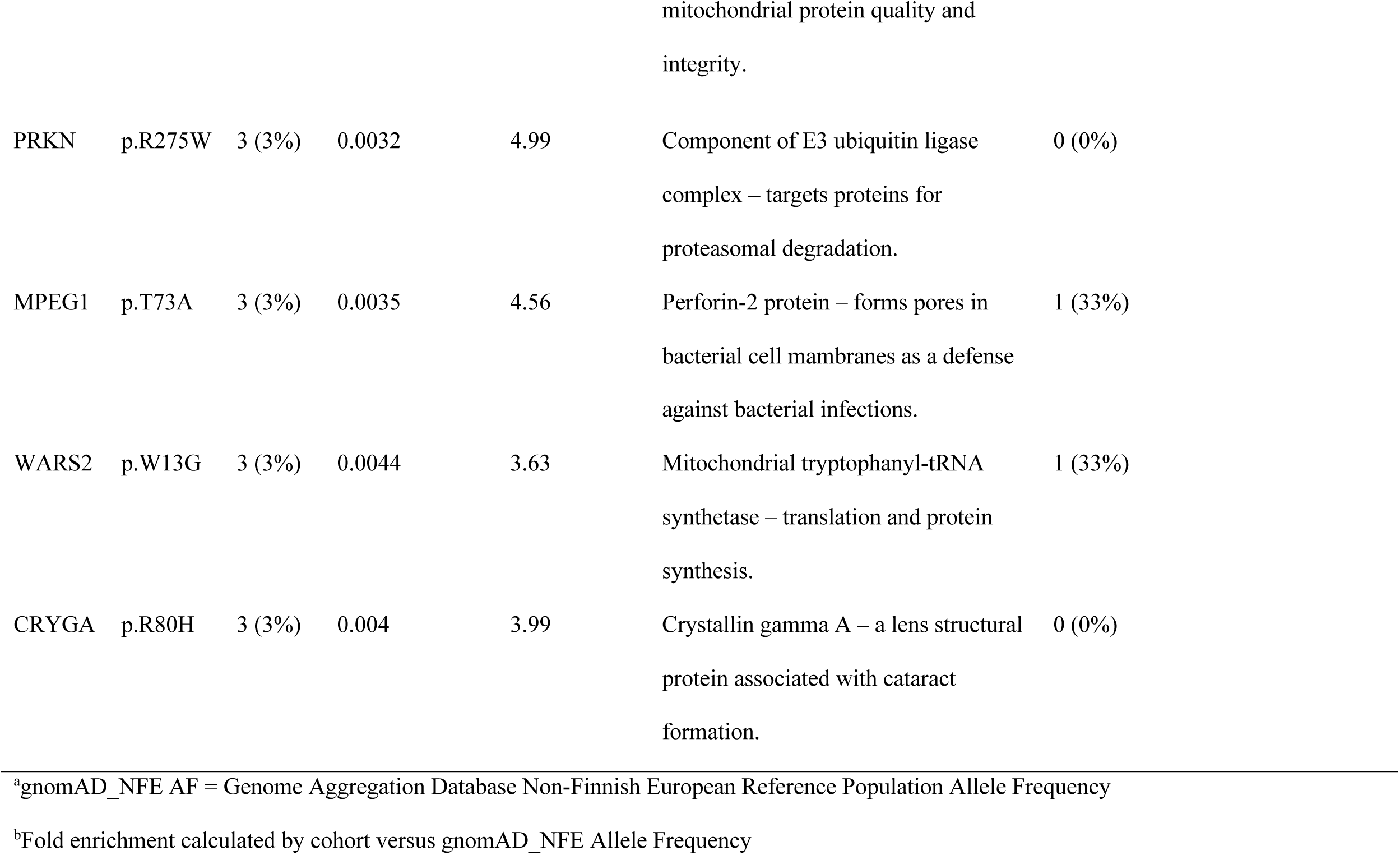
Enriched Germline Pathogenic Recurrent Variants in hfBCC Cohort (N = 94). Table of enriched germline pathogenic recurrent variants in the hfBCC cohort ordered by frequency of recurrence (N or number patients with variant), detailing allele frequency fold enrichment in comparison to reference population database gnomAD_NFE, biologic function of the gene, and number of patients with the variant that had history of internal cancer. gnomAD_NFE AF = Genome Aggregation Database Non-Finnish European Reference Population Allele Frequency. Fold enrichment for each comparison was calculated by dividing cohort variant allele frequency by gnomAD_NFE reference population allele frequency.

**Table 3.** Characteristics of Patients with Germline EPHB2 p.D679N Variant. Table of characteristics of the five patients with hotspot variant *EPHB2* p.D679N. Most carriers developed their first BCC before age 50, younger than the reported average age of first BCC diagnosis in the general population at 63 years old.^37^ Four of five carriers (80%) had a history of an internal malignancy, and of note, all three male carriers had a history of prostate cancer (p=0.024, Fisher’s exact test relative to SEER estimates).

| Patient | Gender | Fitzpatrick | Age of First | Current | # | History of | Family | Family | Family History |
| --- | --- | --- | --- | --- | --- | --- | --- | --- | --- |
|  |  | Skin Type | BCC | Age | BCCs<sup>a</sup> | Internal | History of | History of | of Internal |
|  |  |  | Diagnosis |  |  | Malignancy | NMSC<sup>b</sup> | Melanoma | Malignancy |
| 1 | F | II | 20s | 40s | 14 | No | No | Yes | No |
| 2 | M | II | 30s | 70s | 14 | Prostate cancer | Yes | No | Mother - lung and<br>brain cancers;<br>Father - lung<br>cancer |
| 3 | F | II | 40s | 50s | 8 | Parotid tumor | Yes | No | Father - throat<br>cancer |
| 4 | M | II | 40s | 70s | 21 | Prostate cancer,<br>Hairy cell<br>leukemia | No | No | Father - prostate<br>cancer |
| 5 | M | II | 60s | 70s | 15 | Prostate cancer,<br>Chronic<br>lymphocytic<br>leukemia | Yes | No | No |

<sup>a</sup>Basal cell cancers
<sup>b</sup>Nonmelanoma skin cancer

For conventional BCC, the GWAS meta-analysis effect estimate of β = 0.140 was converted to an allelic odds ratio of 1.15 using OR = e^β^. Relative to gnomAD Non-Finnish European reference allele frequency of 0.0057 (q_0_), the corresponding allele frequency among conventional BCC cases was estimated as follows:

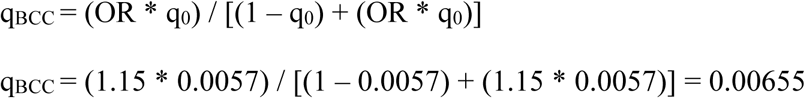

This corresponded to approximately 1.15-fold allele frequency enrichment in conventional BCC.

Relative enrichment was calculated as the ratio of the hfBCC and conventional BCC enrichment estimates:

Relative enrichment = 4.67 / 1.15 = 4.06

Thus, hfBCC ascertainment produced approximately 4-fold greater relative enrichment of p.D679N than conventional BCC ascertainment.

We next performed a post hoc, model-based comparison of the relative case-sample efficiency expected for prospective confirmatory studies using the two ascertainment strategies. Both designs were standardized to a dominant carrier framework, independent case and control groups with 1:1 allocation, 80% statistical power, and a two-sided type I error rate of α = 0.05. Hardy-Weinberg equilibrium was assumed when converting allele ferquencies to carrier probabilities.

The reference carrier probability was calculated as follows:

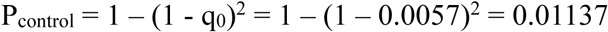

The observed hfBCC carrier frequency was:

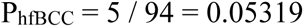

The estimated conventional BCC carrier probability was:

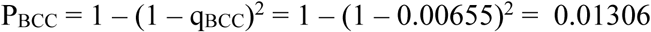

For each ascertainment strategy, model-projected sample sizes for prospective confirmatory studies were estimated using the standard formula for comparing two independent proportions with equal group sizes:

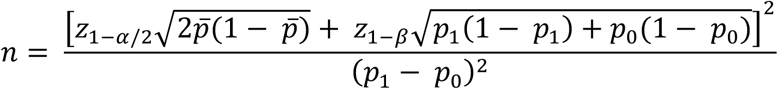

where p_0_ was the reference carrier probability, p_1_ was the carrier probability among cases, *p* = (p_0_ + p_1_) / 2, z_1 – α / 2_ = 1.96, and z_1 – β_ = 0.84. Under these standardized assumptions, projected benchmarks for future prospective confirmatory studies were approximately 280 cases and 280 controls using hfBCC ascertainment and approximately 66,000 cases and 66,000 controls using conventional BCC ascertainment.

Relative case-sample efficiency was defined as the ratio of the model-projected conventional BCC to hfBCC case-sample sizes:

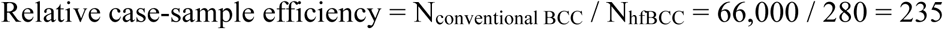

which was reported as an approximately 230-fold estimated improvement in case-sample efficiency with hfBCC ascertainment.

These projections are benchmarks for future prospective confirmatory studies designed to achieve 80% power under standardized assumptions. They do not represent enrollment targets for the present exploratory discovery study. The current 94 patients hfBCC cohort was assembled to identify candidate germline susceptibility variants through extreme-phenotype enrichment, with subsequent population-level and functional evaluation, rather than to provide a stand-alone, prospectively powered confirmation of the p.D679N association. Furthermore, because the hfBCC carrier frequency was estimated in the discovery cohort, the projected sample sizes and relative efficiency estimate may be affected by variant-selection bias and winner’s curse. These results were therefore interpreted as exploratory model-based estimates that require validation in an independently assembled hfBCC cohort.

### Cell culture and transfections

HEK293 human embryonic kidney cells were obtained from ATCC (CRL-1573) (RRID:CVCL_0045) and maintained in Dulbecco’s Modified Eagle’s medium (Cellgro, 10-017-CV) containing fetal bovine serum (Invitrogen, 26140–079) and 1% penicillin-streptomycin (Cellgro, 30-002-CI). The UW-BCC1 cell line was derived from a superficial BCC and is a well-characterized human BCC cell line, featuring Hh and Wnt pathway activation, ability to form tumors with cytological features characteristic of BCC when injected into nude mice, and response to vismodegib.^35^ Human basal cell carcinoma cell lines UW-BCC1 and UW-BCC1-T2 (derived by *in vivo* passaging of the UW-BCC1 cell line in mice)^36^ were provided by Dr. Anthony Oro (Stanford University) and maintained in 154-CF medium containing 7% chelated fetal bovine serum (Invitrogen, 26140–079), 1% penicillin-streptomycin (Cellgro, 30-002-CI) and 0.05mM calcium chloride. All cells were cultured at 37°C and 5% CO_2_.

Transfection of HEK293 and human BCC cell lines with pcDNA3-FLAG tagged wild-type (WT) and D679N EphB2 mutant (MUT) or vector (vec) control were done using Lipofectamine 2000 transfection reagent (ThermoFisher Scientific, 11668027) according to manufacturer’s protocol. A DNA: LF2000 ratio 1:2 or 1:3 was used. Co-transfection of pEGFP-C1 plasmid was done in BCC cell lines to gauge transfection efficiency. Cells were harvested at 48h or 72h, as indicated, for downstream biochemical or functional analysis.

### Plasmids

pcDNA3 (Invitrogen) and pEGFP-C1 (Clontech Laboratories Inc.) plasmids were kindly provided by Dr. Chris Cartwright and Dr. Anson Lowe (Stanford University). Sequence verified pcDNA3 plasmid constructs encoding c-terminal FLAG-tagged WT and G2035A EphB2 (human) were obtained from Genscript (New Jersey, USA).

### Cell lysis and immunoblot analysis

Cells were lysed directly on the plate with cold RIPA buffer (50 mM Tris-HCl, pH 7.6, 100 mM NaCl, 2 mM EDTA,1%Triton X-100 [Sigma, T8787], 0.5% sodium deoxycholate [Sigma, D6750] and 0.1% SDS [Bio-Rad Laboratories, 161–0418]) containing protease inhibitors (Sigma, P8340), 1 mM sodium orthovanadate (Sigma, S6508) and 1 or 10 mM sodium fluoride (Sigma, S1504). The lysates were transferred to microcentrifuge tubes and rotated for 20 min at 4° C. Lysates were cleared by centrifugation at 16,200 x g for 30 min at 4° C and protein estimation was done using Bradford assay reagent (Bio-Rad Laboratories, 500–0205). Equal amount of protein was resolved on 4-12% TGX gels (Bio-Rad Laboratories, Hercules, CA) and analyzed by immunoblotting with appropriate antibodies: EphB2 (clone D2X2I, Cell Signaling Technology, 83029S) (RRID:AB_2800007), FLAG M2 (Sigma, A3320), GAPDH (clone D16H11, Cell Signaling Technology, 5174S) (RRID:AB_10622025), Tubulin (Cell Signaling Technology, 2144S) (RRID:AB_2210548), GFP (B2; Santa Cruz Biotechnology, sc-9996) (RRID:AB_627695), AKT and phospho-Ser473 AKT (Cell Signaling Technology, 9272 and 4060) (RRID:AB_329827; RRID:AB_2315049), Erk and phospho-T202/Y204 Erk (Cell Signaling Technology, 4695S and 4370S) (RRID:AB_390779; RRID:AB_2315112), phosphor-tyrosine (clone 4G10; EMD Millipore, 05-321) (RRID:AB_309678). Primary antibodies were detected using horseradish peroxidase-labeled secondary antibodies (Bio-Rad, NXA931 and NA934) (RRID:AB_772210; RRID:AB_772206) and ECL prime chemiluminescence (Thermo Fisher, 45002401). X-ray film images were scanned at 300 dpi and saved as .tiff images. Densitometry was performed using NIH ImageJ software (RRID:SCR_003070).

### Immunoprecipitations

For immunoprecipitation experiments, transfected cells in 60mm or 10cm tissue culture dishes were washed with PBS (Sigma, D8537) and lysed in plate on ice with 250-400 μl of cold RIPA buffer. Lysates were transferred to 1.5 ml tubes, rotated for 20 min, and clarified by centrifugation at 16,200 x g for 30 min at 4°C. The cleared lysates (1 mg total protein) were diluted 8- to 10-fold with 1% Triton X-100 lysis buffer and incubated with FLAG M2 antibody conjugated to agarose beads (SIGMA, A2220) (RRID:AB_10063035) or FLAG M2 antibody (SIGMA, F3185) (RRID:AB_259529) overnight at 4°C. Twenty μl of protein G agarose beads (EMD Millipore, 16-266) were added (if using unconjugated FLAG M2 antibody) and the tubes rotated at 4°C for 2 h. The beads were washed sequentially with buffers containing 0.1% SDS+1% Triton X-100, 1% Triton X-100 and 1X TBS. Samples were resuspended in 2x Laemmli sample buffer (Bio-Rad Laboratories, 161–0737) containing 2-mercaptoethanol (Sigma, M3148), boiled for 5 min at 95°C and centrifuged briefly. Supernatants were resolved on 4–12% TGX gels (Bio-Rad Laboratories, 456–1084), transferred to PVDF membrane (EMD Millipore, IPVH00010) and analyzed by immunoblotting as described above.

### Biochemical Characterization

48h post transfection of HEK293 cells, cells were stimulated (+) with Ephrin B2-Fc (1.5 mg/ml) for 15 min or left unstimulated (-). Tyrosine phosphorylation of WT and MUT EphB2 was assessed by immunoblotting of FLAG immunoprecipitates with a phospho-tyrosine antibody (4G10) (RRID:AB_309678). Levels of WT and MUT EphB2, phospho-Erk1/2 (T202/Y204) and total Erk levels, and phospho-Akt (Ser473) and total Akt, were evaluated in UW-BCC1 and UW-BCC1-T2 human basal cell carcinoma cells transfected with FLAG-tagged EphB2-WT and MUT or vector control after 48h by immunoblotting of whole cell extract (WCE). Blots and quantitation of EphB2, pAkt, and pErk were normalized to Tubulin and GAPDH, Akt, and Erk, respectively from 2-4 independent experiments.

### Statistical Analysis

Statistical analyses were performed using Prism 10 software (GraphPad, La Jolla, CA) (RRID:SCR_002798). Statistical significance for demographic internal malignancy information and variant enrichment compared to gnomAD reference population was determined using the Chi-squared (χ^2^) test. For SEER internal malignancy analysis, comparisons between observed cancer counts in the hfBCC cohort and expected counts derived from SEER prevalence estimates were performed using Fisher’s exact test for analyses with number hfBCC cancer cases ≤5 and Chi-square test otherwise. For MarketScan^®^ database internal malignancy analysis, logistic regression, adjusting for age and sex, was applied to estimate the associations between BCC groups (BCC 1, 6 or more, and 12 or more, each within 10 years) and control (no BCCs during enrollment) for each cancer. Fisher’s exact test was utilized to determine statistical significance of variant allele frequency of the hfBCC cohort compared to that of control. GWAS meta-analysis was conducted with PLINK (RRID:SCR_001757), which used inverse-variance weighted fixed effects to determine statistical significance. Data in the graphs of Figure 2 are expressed as mean ± standard error of the mean (SEM). Statistical significance was determined using the Student’s unpaired *t* test (two-tailed) when comparing 2 experimental groups. One-way or 2-way ANOVAs and multiple comparison post-tests (Tukey’s honestly significant difference test or Bonferroni test) were used when comparing multiple groups.

## Data Availability

The whole-genome sequencing data will be deposited to the NIH Sequence Read Archive upon publication. Raw 3’-scRNA-seq matrices have previously been deposited in the Gene Expression Omnibus (GEO) database under accession code GSE141526 (RRID:SCR_005012). This study used GWAS summary statistics data from: 1) the *All of Us* Research Program’s Controlled Tier Dataset v7, available to authorized users on the Researcher Workbench, 2) Pan-UK Biobank, GWAS of UK Biobank’s dataset, available to all on Amazon AWS (https://pan.ukbb.broadinstitute.org), and 3) 23andMe, Inc., with data availability restricted under a materials transfer agreement (MTA). The 23andMe GWAS summary statistics data can be provided by 23andMe, Inc. pending scientific review and a completed materials transfer agreement. Requests for this data should be submitted to.

## Code Availability

All scripts and code will be made available on Github upon publication.

## Acknowledgements

The authors thank members of the Sarin and Curtis labs for helpful suggestions and support. We acknowledge Gladstone Histology and Light Microscopy Core Facility for their assistance in conducting immunohistochemistry studies. Certain data were supplied by Merative™ as part of one or more MarketScan^®^ Research Databases. Any analysis, interpretation, or conclusion based on these data is solely that of the authors and not Merative™. MarketScan^®^ data for this project were accessed using the Stanford Center for Population Health Sciences (PHS) Data Core. The PHS Data Core is supported by a National Institutes of Health National Center for Advancing Translational Science Clinical and Translational Science Award (UL1TR003142) and from Internal Stanford funding. The content is solely the responsibility of the authors and does not necessarily represent the official views of the NIH. We gratefully acknowledge *All of Us* participants for their contributions to the *All of Us* GWAS, and thank the National Institutes of Health’s All of Us Research Program for making available GWAS data examined in the GWAS meta-analysis of this study. We would like to thank the research participants and employees of 23andMe, Inc. for making this work possible. Research reported in this publication was supported by funds from the Chan Zuckerberg Biohub Physician-Scientist Fellowship [AC], the Dermatology Foundation [AC, KYS], Stanford Medical Scholars Research Fellowship [VJH, HGC], Howard Hughes Medical Institute Medical Research Fellowship [WC], the American Skin Association [VJH, KYS], the National Cancer Institute of the National Institutes of Health award number K23 CA211793 [KYS], Damon Runyon Cancer Research Foundation Clinical Investigator Award [KYS], and a philanthropic gift by the Swig family [KYS]. This work was supported in part by NIH P30 CA124435 utilizing the Stanford Cancer Institute Shared Resource, Genetics Bioinformatics Service Center. These funders played no role in study design, data collection, analysis and interpretation of data, or the writing of this manuscript.

## Author Contributions

A.C. contributed to conception, study design, data acquisition, whole genome sequencing processing and analysis, SEER and MarketScan cancer prevalence analysis, GWAS meta-analysis, data interpretation, visualization of results, manuscript drafting, and revising, and specifically Figures 1a, 1c, 1d, S2, S3, S4, and Tables 1, 2, 3, and S1, S2, S3, S4, S5. G.S. contributed to conception and experimental design for cell culture transfections and immunoblot analysis, visualization of results, manuscript drafting, and revising, and specifically Figures 2, S5, and S6. G.L. contributed to single-cell RNA sequencing analysis, interpretation, visualization, drafting, and revising, and specifically Figures 1b and S1. M.H. contributed to cell culture transfections and immunoblot analysis, visualization of results, and drafting. H.S. contributed to SEER and MarketScan data acquisition and cancer prevalence analyses, and specifically Tables 1 and S2. S.L. contributed to SEER and MarketScan statistical analysis. V.H. contributed to study conception, design, acquisition of samples, and genetic analysis, and drafting. W.C. contributed to acquisition of samples. H.C. contributed to study conception, design, and acquisition of samples. M.L. contributed to sample acquisition and genetic analysis. K.K. contributed to sample acquisition. J.R. contributed to sample acquisition. H.D. contributed to sample acquisition. M.A. contributed to sample acquisition. I.B. contributed to study design and sample acquisition. 23andMe contributed to data acquisition. K.R. contributed to pathology sample acquisition and visualization. J.H. contributed to data acquisition and genetic analysis. P.U. contributed to reagent acquisition and resources. C.C. contributed to study design, interpretation, and manuscript revising. A.O. contributed to study conception, design, interpretation, and manuscript revising. K.S. contributed to study conception, design, sample and data acquisition, interpretation, validation, visualization, manuscript drafting, and revising. All authors reviewed and approved the final manuscript.

