## SupplementalMaterial for "Leveraging Skin Mutagenesis to Uncover Genetic Cancer Susceptibilities"

### Supplementary Materials:

### Supplementary Figures:

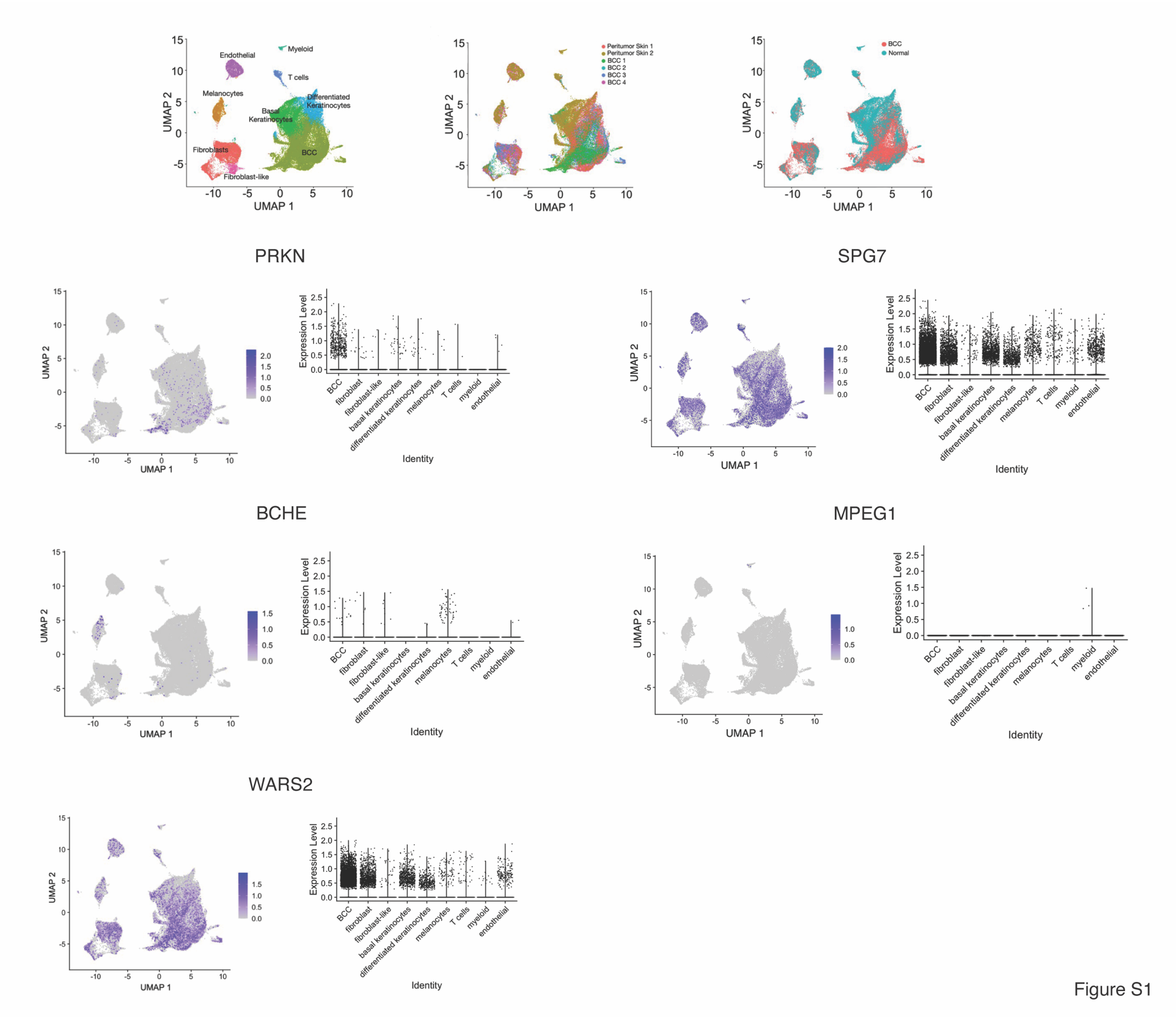


**Figure S1. Single-cell RNA sequencing to assess gene expression within BCC.** Two-dimensional UMAP visualization of single-cell RNA sequencing data obtained from surgical biopsies of basal cell carcinoma (BCC) and matched normal peritumor skin. Referencing **Figure 1b**, single cells were clustered and assigned cell-types according to published annotations,[26] and color-coded according to specific sample types, either BCC or normal peritumor skin. Feature plots highlight individual cells expressing a specific gene of interest, and the corresponding violin plot represents single cells as overlapping dots with each column corresponding to a specific cell type. *PRKN* expression clustered primarily in BCC cells. *BCHE* and *MPEG1* had less expression among BCC cells. *WARS2* and *SPG7* had ubiquitously more expression among multiple cell clusters, including BCC, fibroblast, basal keratinocytes, and differentiated keratinocytes.

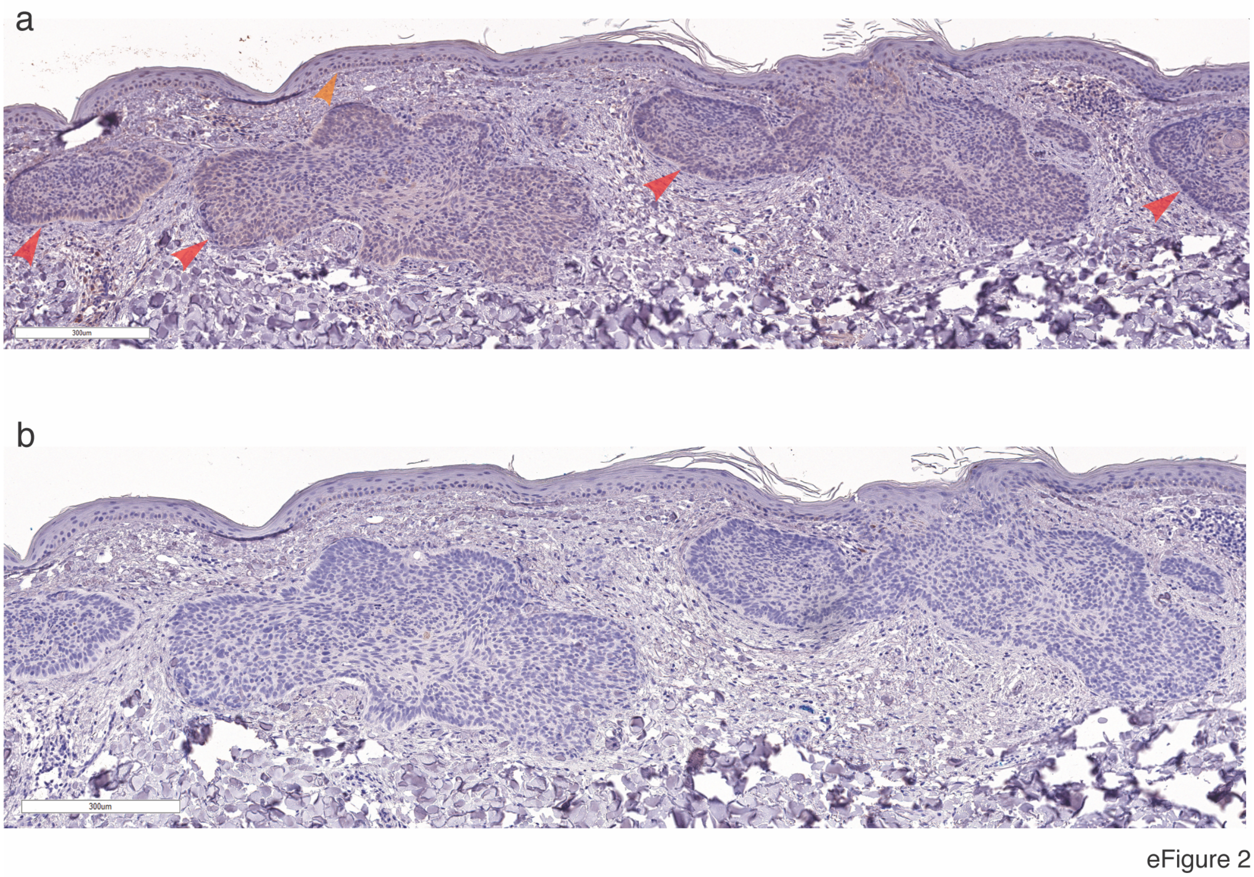


**Figure S2. EphB2 protein level expression within basal cell cancer and epidermal cells.** (**a**) Immunohistochemistry (IHC) of a BCC tumor from a patient with no germline pathogenic variants with EphB2 protein demonstrated staining within basal tumor cells (red arrows) and the epidermis (blue arrow), with minimal to no involvement of the surrounding stroma, demonstrating EphB2 protein level specific expression within keratinocytes, from which BCCs arise. (**b**) In comparison, IHC negative control of the same BCC tumor without primary EphB2 antibody was negative for staining.

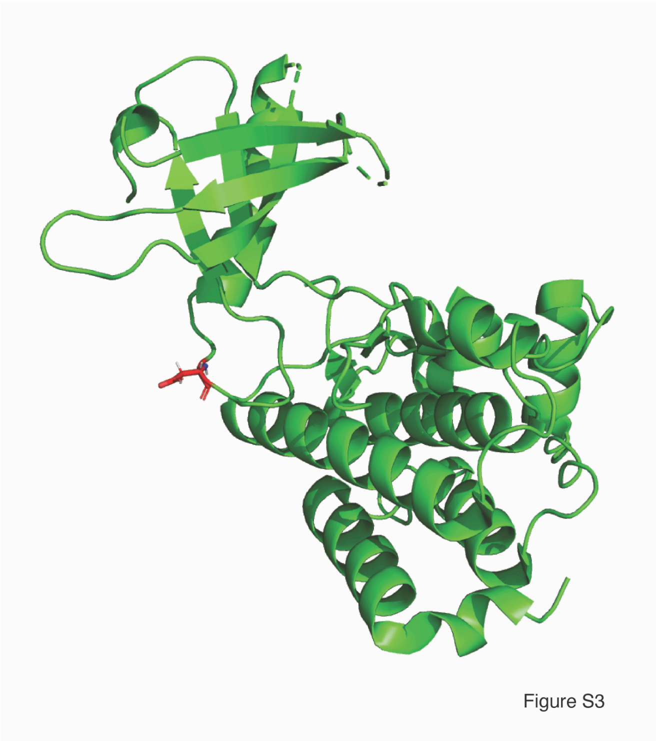


**Figure S3. Ribbon representation of the structure of EphB2 kinase domain** based on Overman, et al. Protein Data Bank (PDB) accession code 3ZFM.[38] Residue D679 (red licorice representation) resides in the kinase domain, linking α helix + β-pleated sheets. This figure was generated using PyMol.

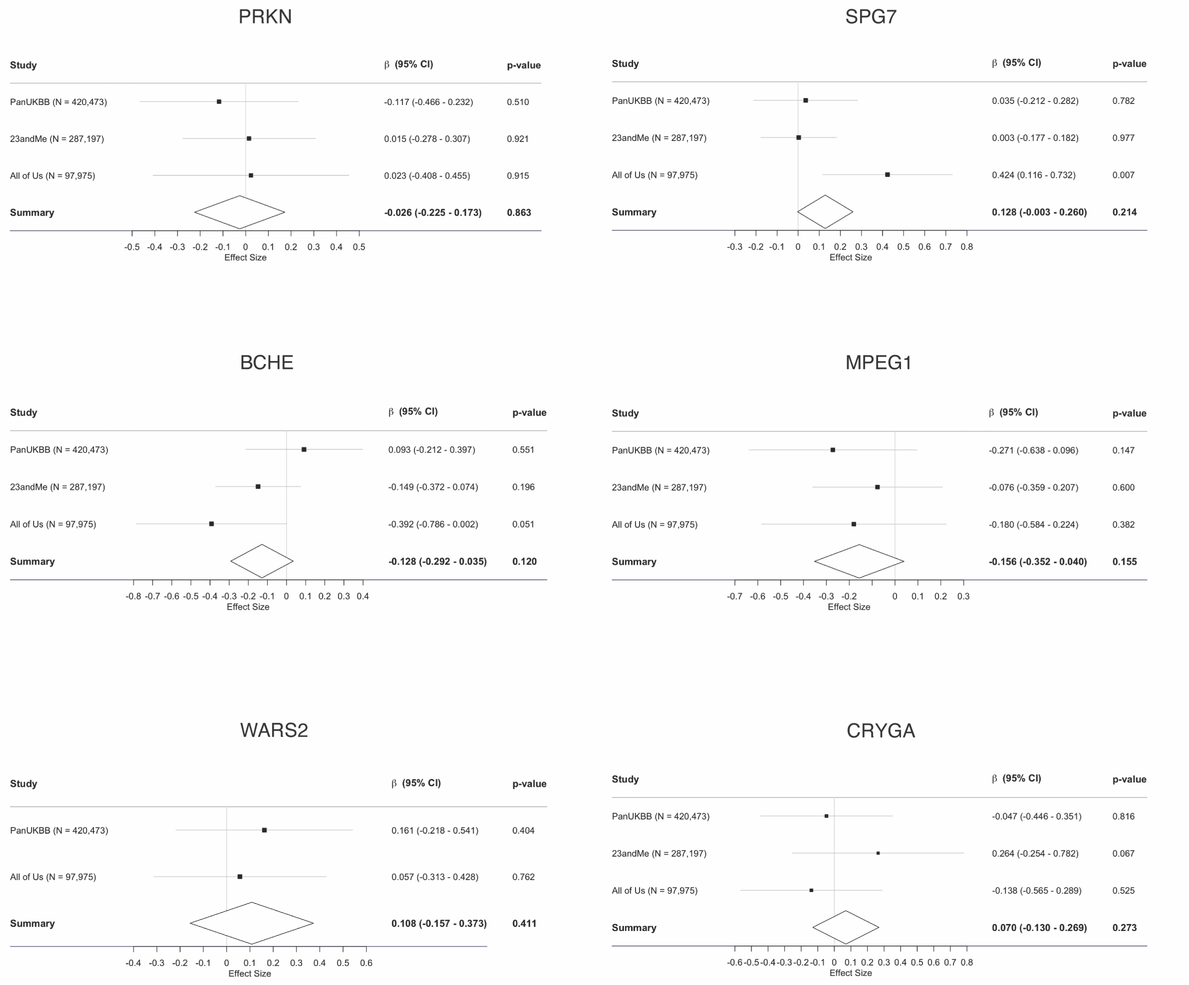


Figure S4

**Figure S4. Forest plot of GWAS meta-analysis of other enriched variants not significant in BCC.** Forest plots of GWAS meta-analyses of enriched variants *BCHE* p.A212V (β = -0.1284, p = 0.1204), *SPG7* p.A510V (β = 0.1284, p = 0.2141), *PRKN* p.R275W (β = -0.0262, p = 0.8631), *MPEG1* p.T73A (β = -0.1564, p = 0.1549), *WARS2* p.W13G (β = 0.1080, p = 0.1314), *CRYGA* p.R80H (β = 0.0696, p = 0.2726) all did not demonstrate significant enrichment in BCC patients (with 1 or more BCCs) within Pan-UK Biobank, 23andMe, and All of Us GWAS cohorts. Error bars = 95% confidence intervals. Diamond = pooled effect.

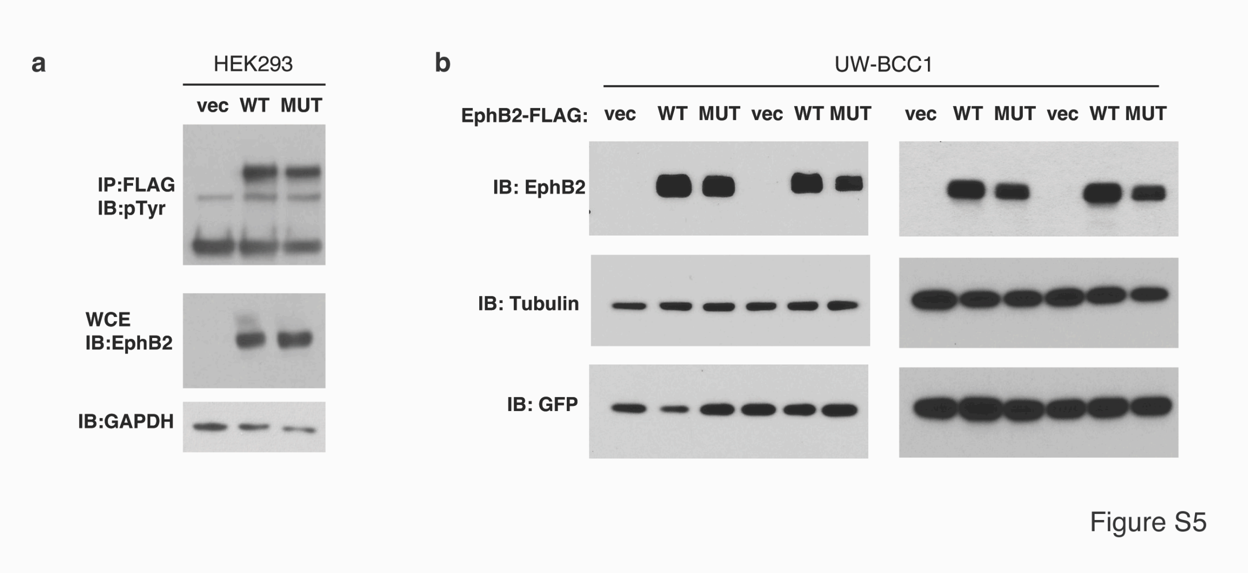


**Figure S5. Related to Figure 2.** (**a**) **Related to Figure 2a.** Basal tyrosine phosphorylation of WT and MUT was evaluated in HEK293 cells transfected with FLAG-tagged WT and MUT EphB2 by immunoprecipitation and immunoblotting with antibodies as indicated, and were similar and reflected total EphB2 levels. (**b**) **Related to Figure 2b.** Comparable transfection efficiency in UW-BCC1 cell line transfected with WT and MUT EphB2 as assessed by co-transfection with EGFP plasmid and immunoblotting of WCE with GFP antibody in experiments where differences in downstream signaling were observed.

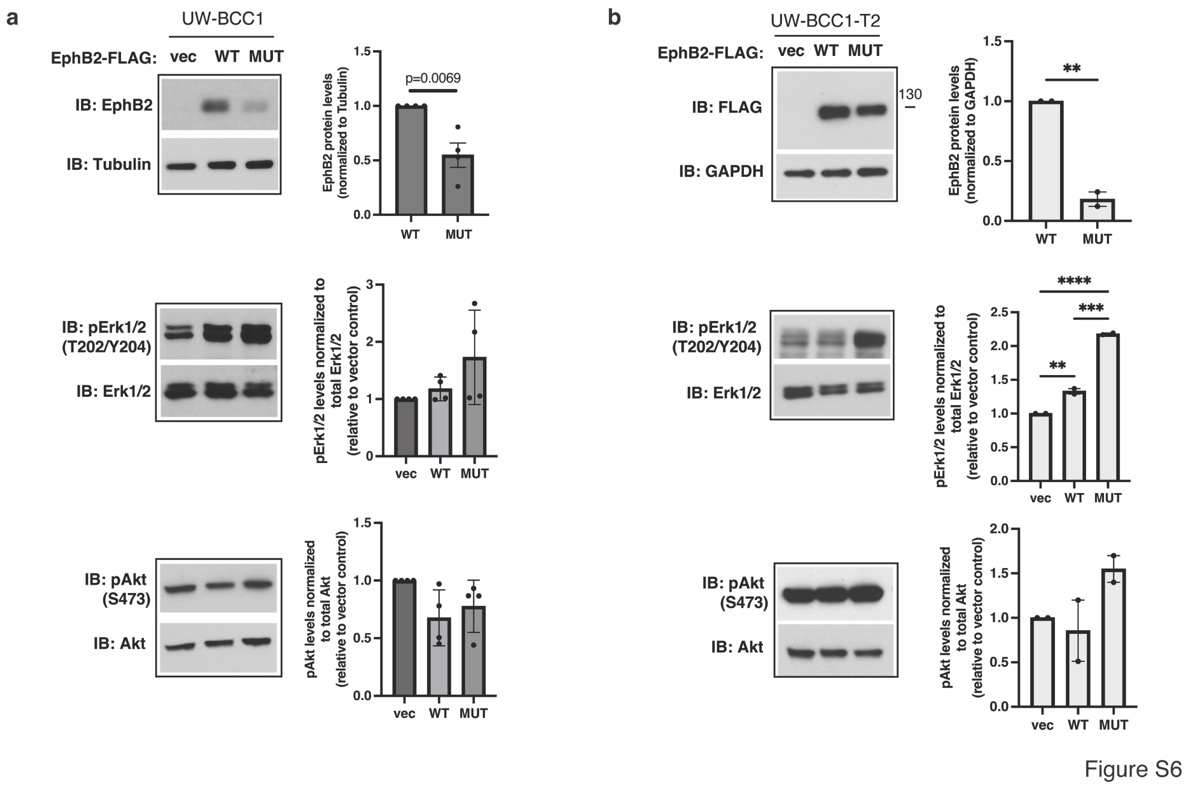


**Figure S6. Cell context dependent differential expression of WT and D679N EphB2 and downstream signaling.** (**a-b**) Levels of WT and MUT EphB2, phosphorylated and total Erk1/2, and phosphorylated and total Akt were evaluated in the (**a**) UW-BCC1 and (**b**) UW-BCC1-T2 human basal cell carcinoma cell lines transfected with EphB2-WT and MUT, by immunoblotting of WCE with antibodies as indicated. MUT EphB2 expression was significantly lower than WT in both cell lines, p = 0.0069 and p = 0.0054, respectively. Upregulation of pErk in MUT compared to WT was significant in UW-BCC1-T2 (p = 0.0003), but not reaching significance in UW-BCC1. Upregulation of pAkt in MUT was also noted in WT, but did not reach significance for either cell line. The differences in downstream mutant effects between cell lines may be due to UW-BCC1-T2 cells possibly gaining enhancement in tumor promoting properties during its derivation by *in vivo* passaging of the parental UW-BCC1 cell line.

**Supplementary Tables:**

**Table S1. hfBCC Cohort Demographics (N = 94).** Table of demographics, characteristics, and oncologic history of hfBCC study patients. SD = standard deviation.

| Characteristics | N | Percent | Mean ± SD (Min-Max) |
|---|---|---|---|
| Gender | | | |
| Male | 59 | 62.8% | |
| Female | 35 | 37.2% | |
| Age at Study Enrollment | | | 66.0 ± 13.7 (28-92) |
| 20-29 | 2 | 2.1% | |
| 30-39 | 2 | 2.1% | |
| 40-49 | 5 | 5.3% | |
| 50-59 | 19 | 20.2% | |
| 60-69 | 19 | 20.2% | |
| 70-79 | 32 | 34.0% | |
| 80-89 | 13 | 13.8% | |
| 90-100 | 2 | 2.1% | |
| Number BCCs in 10 years | | | 15.2 ± 13.7 (6-98) |
| 6-10 | 45 | 47.9% | |
| 11-15 | 24 | 25.5% | |
| 16-20 | 8 | 8.5% | |
| 21-25 | 6 | 6.4% | |
| 26-30 | 5 | 5.3% | |
| 31-35 | 0 | 0.0% | |
| 36-40 | 2 | 2.1% | |
| 41-45 | 1 | 1.1% | |
| 46-50 | 0 | 0.0% | |
| 51-55 | 0 | 0.0% | |
| 56-60 | 1 | 1.1% | |
| ≥61 | 2 | 2.1% | |
| Age at First BCC | | | 40.7 ± 13.6 (18-75) |
| 20-29 | 13 | 15.3% | |
| 30-39 | 27 | 31.8% | |
| 40-49 | 25 | 29.4% | |
| 50-59 | 8 | 9.4% | |
| 60-69 | 7 | 8.2% | |
| 70-79 | 5 | 5.9% | |

| **Characteristics** | **N** | **Percent** | **Mean ± SD (Min-Max)** |
|---|---|---|---|
| Fitzpatrick Skin Type | | | |
| I | 10 | 10.8% | |
| II | 73 | 78.5% | |
| III | 9 | 9.7% | |
| IV | 1 | 1.1% | |
| V | 0 | 0.0% | |
| VI | 0 | 0.0% | |
| Hair Color | | | |
| Red | 9 | 10.1% | |
| Blonde | 38 | 42.7% | |
| Brown | 40 | 44.9% | |
| Black | 2 | 2.2% | |
| Eye Color | | | |
| Blue | 46 | 53.5% | |
| Green | 21 | 24.4% | |
| Brown | 19 | 22.1% | |
| Blistering Sunburns | | | |
| None | 2 | 2.3% | |
| 1-5 | 35 | 39.8% | |
| 6-10 | 9 | 10.2% | |
| ≥11 | 37 | 42.0% | |
| Unsure | 5 | 5.7% | |
| Tanning Bed Use | | | |
| Never | 65 | 73.0% | |
| Once | 3 | 3.4% | |
| ≥2 Times | 21 | 23.6% | |
| Average Hours/Day Outdoors | | | 2.4 ± 2.0 (0-8) |
| 0-1 | 37 | 44.0% | |
| 2-3 | 32 | 38.1% | |
| 4-5 | 5 | 6.0% | |
| 6-7 | 6 | 7.1% | |
| ≥8 | 4 | 4.8% | |

| Characteristics | N | Percent | Mean ± SD (Min-Max) |
|---|---|---|---|
| Sunscreen Use (≥3 days/week) | 39 | 49.4% | |
| Coffee (Cups/Day) | | | 1.7 ± 1.3 (0-6) |
| None | 19 | 21.6% | |
| 1 | 25 | 28.4% | |
| 2 | 23 | 26.1% | |
| 3 | 13 | 14.8% | |
| ≥4 | 8 | 9.1% | |
| Alcohol History | | | 5.0 ± 6.1 (0-40) |
| None/Social/Occasional | 26 | 29.5% | |
| ≤1 drink a day | 45 | 51.1% | |
| 1-2 drinks/day | 12 | 13.6% | |
| 2-3 drinks/day | 4 | 4.5% | |
| >3 drinks/day | 1 | 1.1% | |
| Smoking History | | | |
| Current | 14 | 15.7% | |
| Former | 9 | 10.1% | |
| None | 66 | 74.2% | |
| History of SCC | | | |
| None | 18 | 21.2% | |
| 1-3 | 36 | 42.4% | |
| 4-6 | 12 | 14.1% | |
| 7-10 | 9 | 10.6% | |
| ≥11 | 10 | 11.8% | |
| History of Melanoma | 23 | 25.0% | |
| History of Cutaneous T-cell Lymphoma | 4 | 4.3% | |
| History of Non-Cutaneous Malignancy | 24 | 26.1% | |
| Family History of NMSC[a] | 66 | 72.5% | |
| Family History of Melanoma | 19 | 21.3% | |
| Family History of Non-Cutaneous Malignancy | 49 | 55.1% | |

[a]Nonmelanoma skin cancer

**Table S2. Internal Cancer Prevalence in Patients with a high frequency of BCCs in MarketScan® Database.** Table of cancer prevalence of BCC cohort with BCC 1, 6 or more, and 12 or more within 10 years compared to control patients with no BCCs in the MarketScan® Database. Inclusion criteria were 60 years or older within period 2007-2022 with minimum 10 years enrollment time. Statistical analysis was conducted using logistic regression, adjusting for age and sex, to estimate associations between BCC groups (1, 6 or more, and 12 or more) and control for each cancer. OR = odds ratio. CI = confidence interval.

| Cancer | BCC 1 vs 0 OR (95% CI)[d] | BCC ≥6 vs 0 OR (95% CI)[d] | BCC ≥12 vs 0 OR (95% CI)[d,e] |
|---|---|---|---|
| Head & Neck | 1.316 (1.270 - 1.363)**** | 2.373 (1.999 - 2.818)**** | 3.747 (2.249 - 6.245)**** |
| Gastric | 1.084 (0.989 - 1.188) | 1.871 (1.190 - 2.943)** | 3.032 (0.817 - 11.257) |
| Colorectal | 1.098 (1.059 - 1.138)**** | 1.271 (1.052 - 1.535)* | 1.599 (0.955 - 2.676) |
| Pancreatic | 1.107 (1.016 - 1.206)* | 1.754 (1.012 - 3.043)* | NE |
| Lung | 1.163 (1.113 - 1.215)**** | 1.569 (1.246 - 1.976)*** | 1.781 (0.867 - 3.659) |
| Bone & Soft Tissue | 1.713 (1.608 - 1.824)**** | 5.481 (3.948 - 7.608)**** | 6.288 (2.631 - 15.026)**** |
| Breast[a] | 1.252 (1.217 - 1.287)**** | 1.561 (1.265 - 1.926)**** | 1.751 (0.900 - 3.407) |
| Ovarian[a] | 1.108 (1.022 - 1.201)* | 0.958 (0.538 - 1.705) | 3.050 (0.313 - 29.736) |
| Prostate[b] | 1.075 (1.049 - 1.102)**** | 1.276 (1.141 - 1.426)**** | 1.103 (0.794 - 1.531) |
| Penile[b] | 1.255 (1.084 - 1.453)** | 2.787 (1.300 - 5.978)** | 8.108 (1.011 - 65.041)* |
| Renal | 1.146 (1.084 - 1.211)**** | 1.398 (1.055 - 1.852)* | 2.197 (0.829 - 5.827) |
| Bladder | 1.172 (1.123 - 1.223)**** | 1.415 (1.165 - 1.718)*** | 2.177 (1.233 - 3.844)** |
| Brain | 1.286 (1.181 - 1.400)**** | 2.376 (1.441 - 3.916)*** | NE |
| Thyroid | 1.239 (1.157 - 1.326)**** | 2.011 (1.304 - 3.099)** | 2.513 (0.486 - 13.004) |
| Adrenal | 1.520 (1.279 - 1.806)**** | 4.854 (2.014 - 11.701)*** | NE |
| Any Solid Organ Malignancy | 1.207 (1.189 - 1.224)**** | 1.652 (1.525 - 1.790)**** | 2.150 (1.692 - 2.733)**** |
| Hodgkin's Lymphoma | 1.663 (1.449 - 1.909)**** | 6.042 (2.712 - 13.460)**** | NE |

| | | | |
|---|---|---|---|
| Non-Hodgkin Lymphoma | 1.562 (1.483 - 1.646)**** | 3.035 (2.322 - 3.968)**** | 2.350 (1.109 - 4.980)* |
| Lymphoma | 1.525 (1.452 - 1.601)**** | 3.015 (2.350 - 3.867)**** | 3.640 (1.787 - 7.413)*** |
| Leukemia | 1.593 (1.494 - 1.698)**** | 2.315 (1.705 - 3.143)**** | 4.103 (1.530 - 11.006)** |
| Hematopoietic Malignancy[c] | 1.451 (1.396 - 1.507)**** | 2.540 (2.094 - 3.080)**** | 3.523 (1.991 - 6.232)**** |
| Metastatic Cancer | 1.315 (1.279 - 1.353)**** | 2.065 (1.791 - 2.382)**** | 2.740 (1.801 - 4.168)**** |
| Any Internal Malignancy | 1.278 (1.260 - 1.297)**** | 1.902 (1.759 - 2.056)**** | 2.701 (2.130 - 3.424)**** |

[a]Breast and ovarian cancers analyzed among females.

[b]Prostate and penile cancers analyzed among males.

[c]Hematopoietic malignancy includes Non-Hogkin Lymphoma, Hodgkin's Lymphoma, and leukemia.
[d]OR = Odds Ratios were calculated using logistic regression analysis. Data are adjusted for age at first enrollment and sex.
[e]NE = Not estimable, due to sparse cell counts resulting in model nonconvergence or unstable estimates.

Statistically significant at p < 0.05*, p < 0.01**, p < 0.001***, p < 0.0001****

**Table S3. Enriched Germline Pathogenic Recurrent Variants in hfBCC Cohort – Comparison to Population Databases.** Expanded version of **Table 1** detailing variant rsID, cohort allele frequency, and variant allele frequency fold enrichment in comparison to additional reference population databases ExAC_NFE and 1000g_eur. gnomAD_NFE AF = Genome Aggregation Database Non-Finnish European Reference Population Allele Frequency. ExAC_NFE AF = Exome Aggregation Consortium Non-Finnish European Reference Population Allele Frequency. 1000g_eur AF = 1000 Genomes Project (1000g2015aug_eur) European Reference Population Allele Frequency. Fold enrichment for each comparison was calculated by dividing cohort variant allele frequency by gnomAD_NFE, ExAC_NFE, and 1000g_eur reference population allele frequencies, respectively.

| Gene | Variant | rsID | hfBCC Cohort Allele Frequency (AF) | gnomAD_NFE AF[a] | Fold enrichment vs gnomAD_NFE | ExAC_NFE AF[b] | Fold enrichment vs ExAC_NFE | 1000g_eur AF[c] | Fold enrichment vs 1000g_eur |
|---|---|---|---|---|---|---|---|---|---|
| EPHB2 | p.D679N | rs28936395 | 0.0266 | 0.0057 | 4.67 | 0.0053 | 5.02 | 0.005 | 5.32 |
| BCHE | p.A212V | rs114706984 | 0.0213 | 0.0036 | 5.91 | 0.0029 | 7.34 | 0.002 | 10.64 |
| SPG7 | p.A510V | rs61755320 | 0.0213 | 0.0049 | 4.34 | 0.004 | 5.32 | 0.005 | 4.26 |
| PRKN | p.R275W | rs34424986 | 0.0160 | 0.0032 | 4.99 | 0.0031 | 5.15 | 0.001 | 15.96 |
| MPEG1 | p.T73A | rs143527301 | 0.0160 | 0.0035 | 4.56 | 0.0032 | 4.99 | 0.005 | 3.19 |
| WARS2 | p.W13G | rs139548132 | 0.0160 | 0.0044 | 3.63 | 0.0044 | 3.63 | 0.005 | 3.19 |
| CRYGA | p.R80H | rs139353014 | 0.0160 | 0.004 | 3.99 | 0.0044 | 3.63 | 0.005 | 3.19 |

[a]gnomAD_NFE AF = Genome Aggregation Database Non-Finnish European Reference Population Allele Frequency

[b]ExAC_NFE AF = Exome Aggregation Consortium database Non-Finnish European Reference Population Allele Frequency

[c]1000g_eur AF = 1000 Genomes Project European Reference Population Allele Frequency

**Table S4. ICD-9 and ICD-10 diagnosis codes for identification of cancers in MarketScan® Database.** Table of ICD diagnosis codes to determine BCC count and history of internal cancer.

| Diagnosis | ICD-9 codes (2007-2015) | ICD-10 codes (2015-2022) |
|---|---|---|
| Basal cell carcinoma | 173.01, 173.11, 173.21, 173.31, 173.41, 173.51, 173.61, 173.71, 173.81, 173.91 | C44.01, C44.011, C44.012, C44.111, C44.112, C44.119, C44.211, C44.212, C44.219, C44.311, C44.312, C44.319, C44.41, C44.411, C44.412, C44.511, C44.512, C44.519, C44.611, C44.612, C44.619, C44.711, C44.712, C44.719, C44.81, C44.91 |
| Head, Neck, and Upper Aerodigestive Tract Malignancy | 140.0-149.9, 160.0-161.9, 162.0-163.9, 196.0-196.1, V10.12 | C00.0, C00.1, C00.2, C00.3, C00.4, C00.5, C00.6, C00.8, C00.9, C01, C02.0, C02.1, C02.2, C02.3, C02.4, C02.8, C02.9, C03.0, C03.1, C03.9, C04.0, C04.1, C04.8, C04.9, C05.0, C05.1, C05.2, C05.8, C05.9, C06.0, C06.1, C06.2, C06.8, C06.9, C07, C08.0, C08.1, C08.8, C08.9, C09.0, C09.1, C09.8, C09.9, C10.0, C10.1, C10.2, C10.3, C10.8, C10.9, C11.0, C11.1, C11.2, C11.3, C11.8, C11.9, C12, C13.0, C13.1, C13.2, C13.8, C13.9, C14.0, C14.2, C14.8, C30.0, C30.1, C31.0, C31.1, C31.2, C31.3, C31.8, C31.9, C32.0, C32.1, C32.2, C32.3, C32.8, C32.9, C33, C34.00, C34.01, C34.02, C34.10, C34.11, C34.12, C34.20, C34.21, C34.22, C34.30, C34.31, C34.32, C34.80, C34.81, C34.82, C34.90, C34.91, C34.92, Z85.11, Z85.21, Z85.22 |
| Esophageal Cancer | 150.0-150.9, V10.01 | C15.0, C15.1, C15.2, C15.3, C15.4, C15.5, C15.8, C15.9, Z85.01 |
| Gastric Cancer | 151.0-151.9, V10.02 | C16.0, C16.1, C16.2, C16.3, C16.4, C16.5, C16.6, C16.8, C16.9, Z85.02 |
| Intestinal Cancer | 152.0-152.9, 159.0-159.9, V10.03 | C17.0, C17.1, C17.2, C17.3, C17.8, C17.9, C26.0, Z85.03 |
| Colorectal cancer | 153.0-153.9, 154.0-154.8, V10.04-V10.05, V10.00, V10.09 | C18.0, C18.1, C18.2, C18.3, C18.4, C18.5, C18.6, C18.7, C18.8, C18.9, C19, C20, C21.0, C21.1, C21.2, C21.8, C26.8, C26.9, Z85.04, Z85.05, Z85.09 |
| Liver/biliary cancer | 155.0-155.2, 156.0-156-9, V10.06, V10.08 | C22.0, C22.1, C22.2, C22.3, C22.4, C22.7, C22.8, C22.9, C23, C24.0, C24.1, C24.8, C24.9, Z85.06, Z85.08 |
| Pancreatic cancer | 157.0-157.9, V10.07 | C25.0, C25.1, C25.2, C25.3, C25.4, C25.7, C25.8, C25.9, Z85.07 |
| Spleen cancer | 158.0 | C26.1 |
| Lung cancer | 162.2-162.9, V10.11, V10.19 | C34.00, C34.01, C34.02, C34.10, C34.11, C34.12, C34.20, C34.21, C34.22, C34.30, C34.31, C34.32, C34.80, C34.81, C34.82, C34.90, C34.91, C34.92, C39.0, C39.8, C39.9, Z85.118, Z85.12, Z85.25 |

| Diagnosis | ICD-9 codes (2007-2015) | ICD-10 codes (2015-2022) |
|---|---|---|
| Mediastinum | 163.0-163.9, 164.0-164.9, V10.13, V10.14 | C37, C38.0, C38.1, C38.2, C38.3, C38.4, C38.8, Z85.13, Z85.14, Z85.23, Z85.24 |
| Bone/Soft Tissue | 170.0-170.9, 171.0-171.9 | C40.00, C40.01, C40.02, C40.10, C40.11, C40.12, C40.20, C40.21, C40.22, C40.30, C40.31, C40.32, C40.80, C40.81, C40.82, C40.90, C40.91, C40.92, C41.0, C41.1, C41.2, C41.3, C41.4, C41.8, C41.9, C49.0, C49.1, C49.2, C49.3, C49.4, C49.5, C49.6, C49.8, C49.9 |
| Breast cancer | 174.0-174.9, 175.0-175.9, V10.3 | C50.011, C50.012, C50.019, C50.021, C50.022, C50.029, C50.111-C50.119, C50.121-C50.129, C50.211-C50.219, C50.221-C50.229, C50.311-C50.319, C50.321-C50.329, C50.411-C50.419, C50.421-C50.429, C50.511-C50.519, C50.521-C50.529, C50.611-C50.619, C50.621-C50.629, C50.811-C50.819, C50.821-C50.829, C50.911-C50.919, C50.921-C50.929, Z85.3 |
| Vulvar cancer | 184, V10.44 | C51.0, C51.1, C51.2, C51.8, C51.9, Z85.44 |
| Cervical cancer | 180.0-180.9, V10.41 | C52, C53.0, C53.1, C53.8, C53.9, Z85.41 |
| Uterine cancer | 179, 182.0-182.9, V10.42 | C54.0, C54.1, C54.2, C54.3, C54.8, C54.9, C55, Z85.42 |
| Ovarian Cancer (and other female genital) | 183.0, 183.2-183.9, V10.40, V10.43 | C56.1, C56.2, C56.9, C57.00, C57.01, C57.02, C57.10, C57.11, C57.12, C57.20, C57.21, C57.22, C57.3, C57.4, C57.7, C57.8, C57.9, C58, Z85.43 |
| Penile cancer | 187.1-187.9, V10.47 | C60.0, C60.1, C60.2, C60.8, C60.9, Z85.47 |
| Prostate cancer | 185, V10.45 | C61, Z85.45 |
| Testicular cancer | 186.0-186.9, V10.46 | C62.00, C62.01, C62.02, C62.10, C62.11, C62.12, C62.90, C62.91, C62.92, C63.0, C63.1, C63.2, C63.7, C63.8, C63.9, Z85.46 |
| Renal cancer | 189.0-189.1, V10.49 | C64.1, C64.2, C64.9, C65.1, C65.2, C65.9, Z85.49 |
| Bladder/Urinary cancer | 188.0-188.9, 189.2-189.4, 189.8-189.9, V10.48 | C66.1, C66.2, C66.9, C67.0, C67.1, C67.2, C67.3, C67.4, C67.5, C67.6, C67.7, C67.8, C67.9, C68.0, C68.1, C68.8, C68.9, Z85.51, Z85.48 |
| Eye cancer | 190.0-190.9 | C69.00, C69.01, C69.02, C69.10, C69.11, C69.12, C69.20, C69.21, C69.22, C69.30, C69.31, C69.32, C69.40, C69.41, C69.42, C69.50, C69.51, C69.52, C69.60, C69.61, C69.62, C69.80, C69.81, C69.82, C69.90, C69.91, C69.92, Z85.840 |

| Diagnosis | ICD-9 codes (2007-2015) | ICD-10 codes (2015-2022) |
|---|---|---|
| Brain cancer | 191.0-191.9, 192.0-192.9, V10.85 | C70.0, C70.1, C70.9, C71.0, C71.1, C71.2, C71.3, C71.4, C71.5, C71.6, C71.7, C71.8, C71.9, C72.0, C72.1, C72.20, C72.21, C72.22, C72.30, C72.31, C72.32, C72.40, C72.41, C72.42, C72.50, C72.51, C72.52, C72.59, C72.8, C72.9, Z85.841, Z85.842, Z85.848, Z85.840 |
| Thyroid cancer | 193, V10.87 | C73, Z85.850 |
| Adrenal cancer | 194.0-194.1 | C74.00, C74.01, C74.02, C74.10, C74.11, C74.12, C74.90, C74.91, C74.92, Z85.858 |
| Other Endocrine cancer (parathyroid, pituitary, pineal, etc.) | 194.3-194.9 | C75.0, C75.1, C75.2, C75.3, C75.4, C75.5, C75.9, Z85.859 |
| Hodgkin's lymphoma | 201.0-201.9, V10.71 | C81.00, C81.01, C81.02, C81.03, C81.04, C81.05, C81.06, C81.07, C81.08, C81.09, C81.10, C81.11, C81.12, C81.13, C81.14, C81.15, C81.16, C81.17, C81.18, C81.19, C81.20, C81.21, C81.22, C81.23, C81.24, C81.25, C81.26, C81.27, C81.28, C81.29, C81.30, C81.31, C81.32, C81.33, C81.34, C81.35, C81.36, C81.37, C81.38, C81.39, C81.40, C81.41, C81.42, C81.43, C81.44, C81.45, C81.46, C81.47, C81.48, C81.49, C81.70, C81.71, C81.72, C81.73, C81.74, C81.75, C81.76, C81.77, C81.78, C81.79, C81.90, C81.91, C81.92, C81.93, C81.94, C81.95, C81.96, C81.97, C81.98, C81.99, Z85.71 |
| Non-hodgkin's lymphoma (and other malignant lymphoid neoplasms) | 200.0-200.8, 202.0-202.9, V10.72 | C82.00-C82.69, C82.80-C82.99, C83.00-C83.19, C83.30-C83.39, C83.50-C83.59, C83.70-C83.99, C84.00-C84.19, C84.40-C84.49, C84.60-C84.99, C85.10-C85.29, C85.80-C85.99, C86.0-C86.6, C88.0, C88.3, C88.8, C88.9, C96.2, C96.3, C96.8, C96.9, Z85.72 |
| Multiple myeloma/Plasma-cell neoplasms | 203.0-203.1 | C90.0-C90.02, C90.10, C90.12, C90.20, C90.22, C90.30, C90.32, Z85.79 |
| Acute Lymphoblastic Leukemia (ALL) | 204.0 | C91.00-C91.02 |
| Acute myeloid leukemia (AML) | 205.0 | C92.00-C92.02 |

| Diagnosis | ICD-9 codes (2007-2015) | ICD-10 codes (2015-2022) |
|---|---|---|
| Other Acute Leukemias (APL, AMoL, unspecified, subacute) | 206.0, 207.0, 208.0, 208.2 | C92.40-C92.42, C93.00-C93.02, C94.00-C94.02, C94.20-C94.22, C95.00-C95.02, C95.20-C95.22 |
| Chronic Lymphocytic Leukemia (CLL) | 204.1 | C91.10-C91.12 |
| Chronic myeloid leukemia (CML) | 205.1 | C92.10-C92.12 |
| Other Chronic Leukemias (Hairy cell leukemia, atypical CML, CMML, unspecified) | 206.1, 207.1, 208.1 | C91.40-C91.42, C91.50-C91.52, C92.20-C92.22, C93.10-C93.12, C94.30-C94.32, C95.10-C95.12 |
| Other Leukemias (mixed phenotype, monocytic leukemias, unspecified) | 206.9, 208.8, 208.9 | C93.30-C93.32, C93.90-C93.92, C94.40-C94.42, C94.80-C94.82, C95.90-C95.92 |
| History of Lymphoma | V10.79 | Z85.79 |
| History of Leukemia | V10.6, V10.60, V10.61, V10.62, V10.63, V10.69 | Z85.6, Z85.60, Z85.61, Z85.62, Z85.63, Z85.69 |
| History of other malignancy | V10.89, V10.9 | Z85.89, Z85.9 |
| Metastatic cancer | 196.0-196.9, 197.0-197.8, 198.0-198.89, 199.0-199.1 | C77.0, C77.1, C77.2, C77.3, C77.4, C77.5, C77.8, C77.9, C78.00, C78.01, C78.02, C78.1, C78.2, C78.30, C78.39, C78.40, C78.41, C78.42, C78.43, C78.49, C78.5, C78.6, C78.7, C78.80, C78.89, C79.00, C79.01, C79.02, C79.10, C79.11, C79.12, C79.19, C79.2, C79.31, C79.32, C79.40, C79.49, C79.51, C79.52, C79.60, C79.61, C79.62, C79.63, C79.70, C79.71, C79.72, C79.81, C79.82, C79.89, C79.9, C80.0, C80.1, C80.2 |

**Table S5. CPT procedure codes utilized for association with basal cell carcinoma in MarketScan® Database.** Table of CPT procedure codes to associate with ICD diagnosis codes to determine BCC count.

| **Procedure** | **CPT codes (2007-2022)** |
|---|---|
| Destruction procedure | 17260, 17261, 17262, 17263, 17264, 17266, 17270, 17271, 17272, 17273, 17274, 17276, 17280, 17281, 17282, 17283, 17284, 17286 |
| Excision procedure | 11600, 11601, 11602, 11603, 11604, 11606, 11620, 11621, 11622, 11623, 11624, 11626, 11640, 11641, 11642, 11643, 11644, 11646, 67840, 67961, 67966 |
| Mohs procedure | 17311, 17313 |
